# Deep learning system for brain image-aided diagnosis of multiple major mental disorders

**DOI:** 10.1101/2022.06.01.22275855

**Authors:** Qingfeng Li, Wengzheng Wang, Qian Guo, Lijuan Jiang, Kaini Qiao, Yang Hu, Xiaochen Zhang, Zhen Wang, Daihui Peng, Qing Fan, Min Zhao, Yiru Fang, Jijun Wang, Hong Qiu, Jinhong Wang, Guanjun Li, Jianhua Sheng, Chunbo Li, Zhi Yang, the Psychiatric Imaging Consortium

## Abstract

The current clinical diagnosis of psychiatric disorders relies heavily on subjective assessment of symptoms. While neuroimaging has made an essential contribution to characterizing the brain of psychiatric disorders, it does not currently serve the clinical diagnosis of major psychiatric disorders. Here, we report a neuroimaging-aided diagnostic system for major psychiatric disorders designed for clinical needs. We developed novel deep learning networks with attentional mechanisms and applied them to a large-scale, single-center neuroimaging dataset containing four major psychiatric disorders and healthy groups (n=2490). Both cross-validation and extensive independent validation using multiple open-source datasets (n = 1972) showed that the system could accurately identify any one of the four diagnostic categories and healthy population from brain structural imaging. For the first time, we have constructed an automatic neuroimaging-aid diagnostic system that considers common issues in practice, such as co-morbid diagnoses and the discrimination between specific suspected diagnoses. Furthermore, real-world applications have validated the system’s effectiveness. These works contribute to the translation of brain research to objective diagnostic aids for psychiatric disorders.

## Introduction

Mental disorders are diseases of the human brain that can cause suffering to the patient or others, severely impair social functioning, and have become the largest economic burden of disease in the world^1^. Bipolar disorder (BD), major depressive disorder (MDD), schizophrenia (SZ), and obsessive-compulsive disorder (OCD) are four clinically common and severe adult psychiatric disorders^2^. Despite significant advances in neurobiological research on these psychiatric disorders, clinical diagnosis still relies on subjective evaluation of behavioral symptoms^3^. The complexity of symptoms^4–6^ and the subjectivity of assessment methods impede accurate and consistent diagnosis. Researchers are striving to find objective techniques to assist in the diagnosis of these psychiatric disorders or subtypes of the disease.

As a non-invasive, high-throughput, and widely available measure, T1-weighted MRI can reflect fine-grained features of brain structure^7–9^ that have been associated with environmental and genetic factors for mental disorders^10–20^. Combining machine learning and T1-weighted images to identify certain neurodegenerative diseases, such as Alzheimer’s disease and mild cognitive impairment, has seen some success^21–27^, supporting the potential to application of this approach as an objective aid for diagnosing major severe psychiatric disorders. Despite its importance for the development of psychiatric diagnosis as a more objective discipline, there is still a lack of neuroimaging-aided diagnostic systems that span a variety of major psychiatric disorders and meet the practical needs of clinical diagnosis.

In clinical practice, the greatest difficulty psychiatrists encounter is not in identifying a diagnostic category in healthy individuals. Instead, they are challenged to identify a person’s primary diagnosis from among multiple possible diagnoses, based on a symptom set with considerable individual variation. This is important for the adoption of effective intervention strategies. For example, the depressive episodes of BD may have similar symptoms to those of MDD, but the treatment strategies for the two disorders are different. The wrong treatment strategy may instead exacerbate symptoms. Meanwhile, because symptom overlap and comorbidity are common in the real world^4, 6, 28^, accurate identification of co-morbidity in clinical practice can be difficult.

Current neuroimaging studies do not yet meet these practical needs. Previous attempts have mainly focused on distinguishing certain types of diagnosis from health control^27, 29–32^. Some studies have built classifiers to distinguish between two or three diagnoses^33–39^, but the sensitivity and specificity of these classifiers have only been tested in two categories and not in the context of multiple diagnostic categories. Neuroimaging-assisted diagnostic systems that infer the possibility of multiple diagnostic types simultaneously and suggest possible co-morbid patterns are lacking.

Furthermore, most classifiers have been validated only using “clean” samples that were collected following strict inclusion and exclusion criteria for scientific research. While the “clean” samples help reduce heterogeneity, they may not reflect the real difficulties in clinical practice. Due to the sophistication of mental illness and the wide range of individual differences, even for an experienced psychiatrist, making a definitive diagnosis at a patent’s first visit is not an easy task. Therefore, whether classifiers trained with scientific data can be used for diagnosis aids in the real world warrant further evaluations.

Here, we construct a neuroimaging-assisted diagnostic system for major psychiatric disorders to meet clinical needs. To achieve this goal, we developed a new spatial patches-based hierarchical deep learning network (PHN) and applied it to a large-scale neuroimaging dataset covering four diagnostic categories and healthy population (n = 2490, SZ, MDD, BD, OCD, HC) to train classifiers to infer multiple diagnostic types based on T1-weighted MRI brain images. The PHN addresses the methodological challenges through a novel two stage architecture to address the methodological challenges, which applies deep learning methods to localize brain regions of interest and extract image features. Beyond pair-wise classifiers, we obtained classifiers that identify a certain type among multiple diagnostic types. Based on these classifiers, we constructed a closed-loop, automatic neuroimaging-assisted diagnostic system connecting a physician workstation, an MRI workstation, a data analysis server, and a physician query and feedback system, and tested the performance of the system in real-world applications. We found that the PHN classifier can accurately identify primary diagnoses of mental disorders and outperforms common machine learning and deep learning models. Key components of the PHN play essential roles in performance. We validated the performance of the PHN classifier using independent large sample data and verified the ability of the neuroimaging-assisted diagnostic system to identify primary diagnoses and suggest co-morbidities based on consensus diagnoses of psychiatrists in real-world applications.

## Results

### A patch-based hierarchical network for image classification (PHN)

For accurate recognizing psychiatric diseases, we designed a patch-based hierarchical neural network (PHN) that augmented with a pre-attention stage (PHN-stage1) and local feature extraction and classification stage (PHN-stage2) (Figure 1). PHN-stage1 pre-locates possible disease-related brain regions to reduce redundant information for PHN-stage2. The core engine of PHN is a patch-based feature extraction network (P-FEN), which extracts diagnosis-sensitive features and evaluates disease probability from the input image patch. Unlike the commonly used convolutional neural networks (CNNs), P-FEN features stacked U-structures for latent feature extraction. Such a design is based on the current state-of-the-art natural image segmentation model^40^, allowing extensive multi-scale features extraction at relatively low computation and memory costs. Specifically, P-FEN is formed by five U-blocks that contain convolutional layers, batch-normalization layers, ReLU layers, and max-pooling layers (if necessary). The feature vectors derived from the global max-pooling layer are fed into a 1-layer perceptron for feature fusion, followed by a soft-max process to generalize normalized probability values. The final prediction probabilities of a disease label are then generated. The detailed design of the P-FEN is described in Supplementary Table S1.

**Figure 1.**
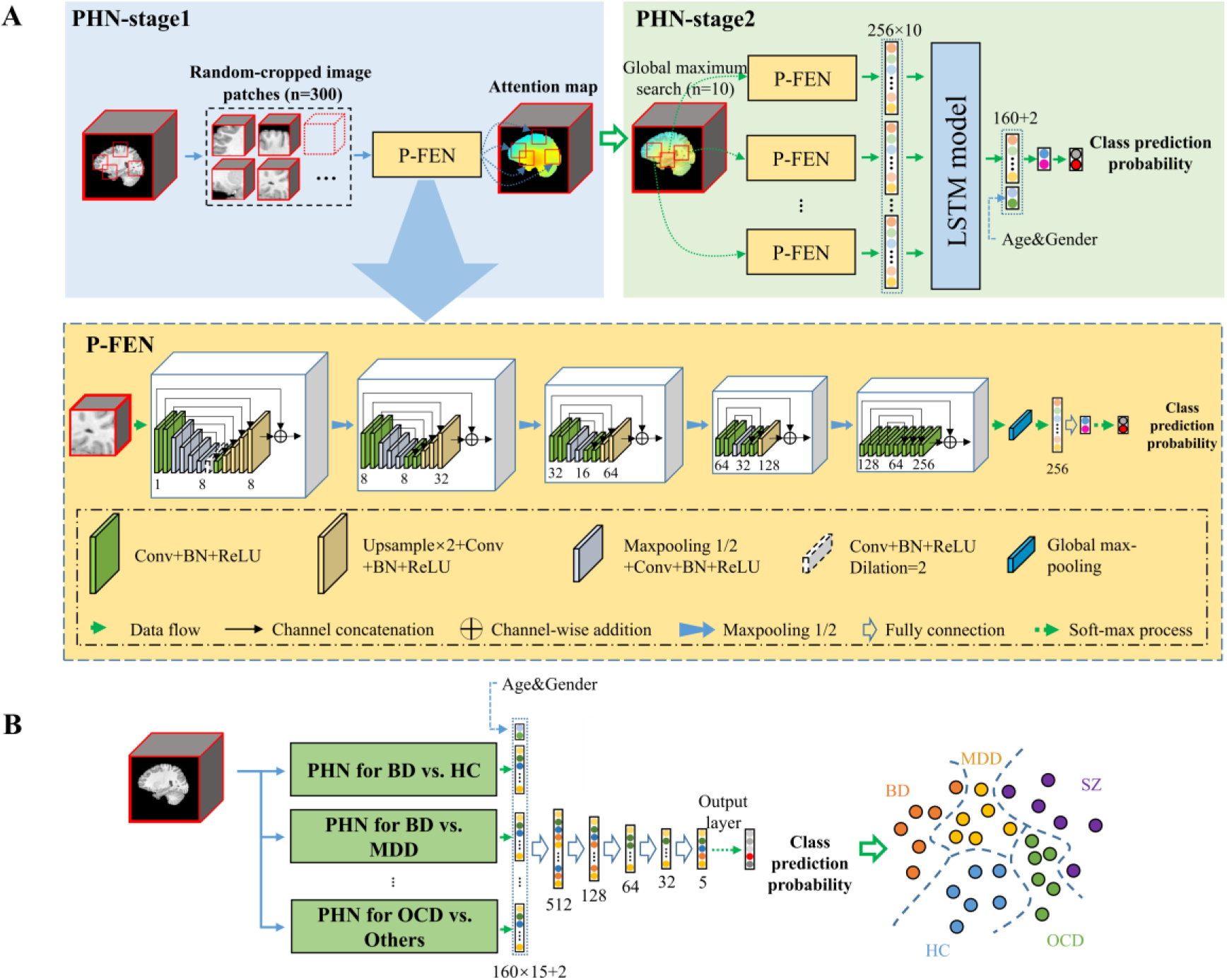
(A) The workflow of PHN. PHN-stage1 randomly samples image patches and input them to the P-FEN. The outputs of P-FEN form an attention map. PHN-stage2 focuses on the image patches with the top confidence and applies P-FEN and LSTM models to fuse the features of the top 10 spatially separated patches to make a final prediction. The detailed structure of the P-FEN, which is the core engine of the PHN, is also shown in (A). P-FEN accepts a 3-dimensional image block as input and obtains the disease probability prediction after several feature extraction filters such as convolutional and pooling layers. The numbers in each U-block are the number of output channels of the input layer, output layer, and hidden layer. (B) Model structure of the PHN multi-disease classifier.

PHN-stage1 randomly extracts 300 image patches with a size of 48×48×48 from the input image and feeds them into the trained P-FEN model to obtain disease probabilities. The centers of the patches are constrained within the brain. Based on the predicted probabilities of the 300 patches, an individual-specific confidence map (i.e., the attention map) is generated (Methods and Supplementary Figure S1). According to the attention map, PHN-stage2 selects ten image patches centered on voxels with peak disease prediction confidence. The algorithm only selects image patches that do not cover the center of each other to avoid information redundancy. The selected image patches are ranked by their prediction confidence and input to the P-FEN model. The feature vectors derived from the P-FENs (behind the global max-pooling layer), are concatenated into a 256×10 matrix. The two dimensions of the matrix reflect local features in a given patch and the order of the spatially separated patches, respectively. This information is then processed by a long-short-term memory (LSTM) model^41^, which considers the inter-dependence of the spatially separated patches as a sequence. The LSTM cell in the model has two hidden layers with a dropout rate of 0.5, and each layer has 16 neurons. LSTM generates a 160-dim feature vector that combines local features of multiple locations in the brain. Age and gender are further concatenated into the output of LSTM, and the final disease prediction probability is derived through a 1-layer perceptron. In order to unify the scale of age and gender with that of the features from LSTM, the value of age is divided by 100, and the gender is set to 0.4 for males and 0.6 for females.

### PHN classifiers accurately recognize one diagnosis from the others

Based on PHN, we trained and tested a serial of image classifiers using T1-weighted 3D brain structural images from 2490 individuals, including 468 BD, 169 MDD, 1285 SZ, 268 OCD, and 300 HC. The data we used were from the Shanghai Mental Health Center Psychiatric Imaging Consortium (PIC), which was contributed by over 10 research groups with high-quality, carefully labeled neuroimaging data that acquired on the same scanner with minimal scan parameter differences (see Methods). Figure 2 presents the performance of PHN classifiers in five-fold cross-validation. The PHN classifiers achieved≥0.90 of the area under the ROC curve (AUC) in 7 of 10 pair-wise classification tasks (BD-HC, BD-MDD, BD-OCD, SZ-HC, SZ-MDD, SZ-OCD, and OCD-HC)(Figure 2A). The lowest AUC appeared in classifying BD and SZ (0.68). More useful classification tasks for clinical practice are recognizing a specific diagnosis type from the others. PHN classifiers achieved 0.90, 0.73, 0.91, 0.84, and 0.93, respectively, for recognizing HC, BD, MDD, SZ, and OCD individuals (Figure 2B). When comparing to other classification algorithms, the PHN classifiers showed higher AUC in all the 15 tasks (Figure 3, Supplementary Table S2).

**Figure 2.**
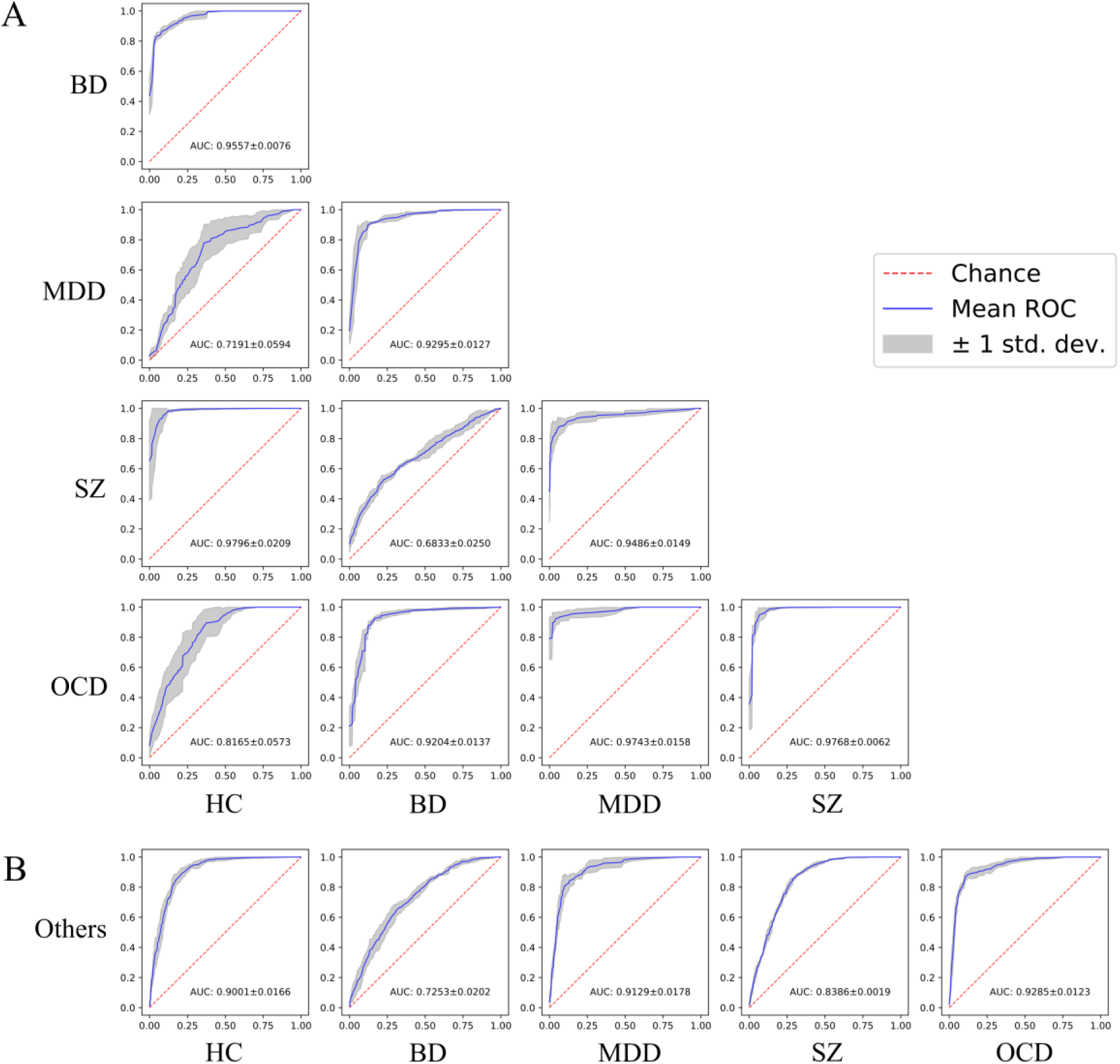
ROC curves of PHN for classifying HC, BD, MDD, SZ, and OCD. (A) ROC curves for classifying all possible pairs of diagnoses. The blue curves represent the averaged ROC, and the grey bands represent the standard deviation of the ROC curves among the five-fold cross-validations. (B) ROC curves of PHN for recognizing specific diagnoses from the others. AUC: area under the ROC curve, presented as mean ± standard deviation.

**Figure 3.**
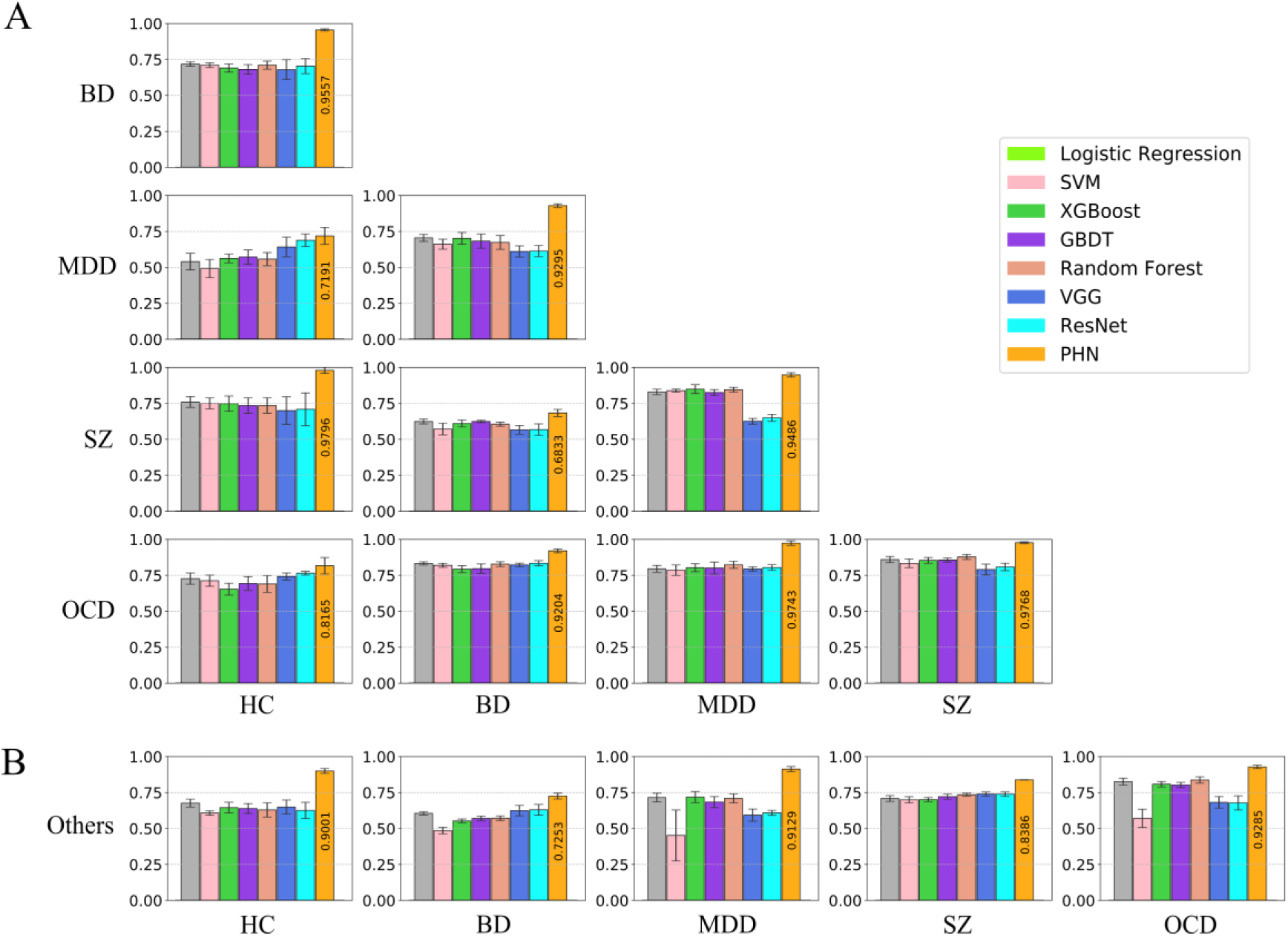
Comparisons of the area under the ROC curve (AUC) between PHN and the other algorithms. (A) All possible pairs of diagnoses; (B) Recognizing specific diagnoses from the others. SVM: support vector machine; XGBoost: extreme gradient boosting, GBDT: gradient boosting decision tree; VGG: 3D visual geometry group network; ResNet: 3D residual network; PHN: patch-based hierarchical network

### The structure of PHN is essential for accurate prediction

We evaluated the effectiveness and robustness of PHN design via a series of ablation studies based on the HC vs. Others task. First, replacing the PHN-stage1 as a random patch selection module reduced the performance of the classifier, as reflected by lower AUC (Figure 4A), supporting the effectiveness of the pre-attention stage. Second, replacing the P-FEN module using other commonly used neural network models, such as VGG and ResNet, degraded the classification performance (Figure 4B), supporting the essential role of P-FEN. Third, replacing the LSTM model in PHN-stage2 with a multi-layer perceptron (PHN_linear in Supplementary Table S2) caused drop of AUC on 12 of all 15 classification tasks (Supplementary Table S2). These findings confirm the designated roles of the modules in PHN.

**Figure 4.**
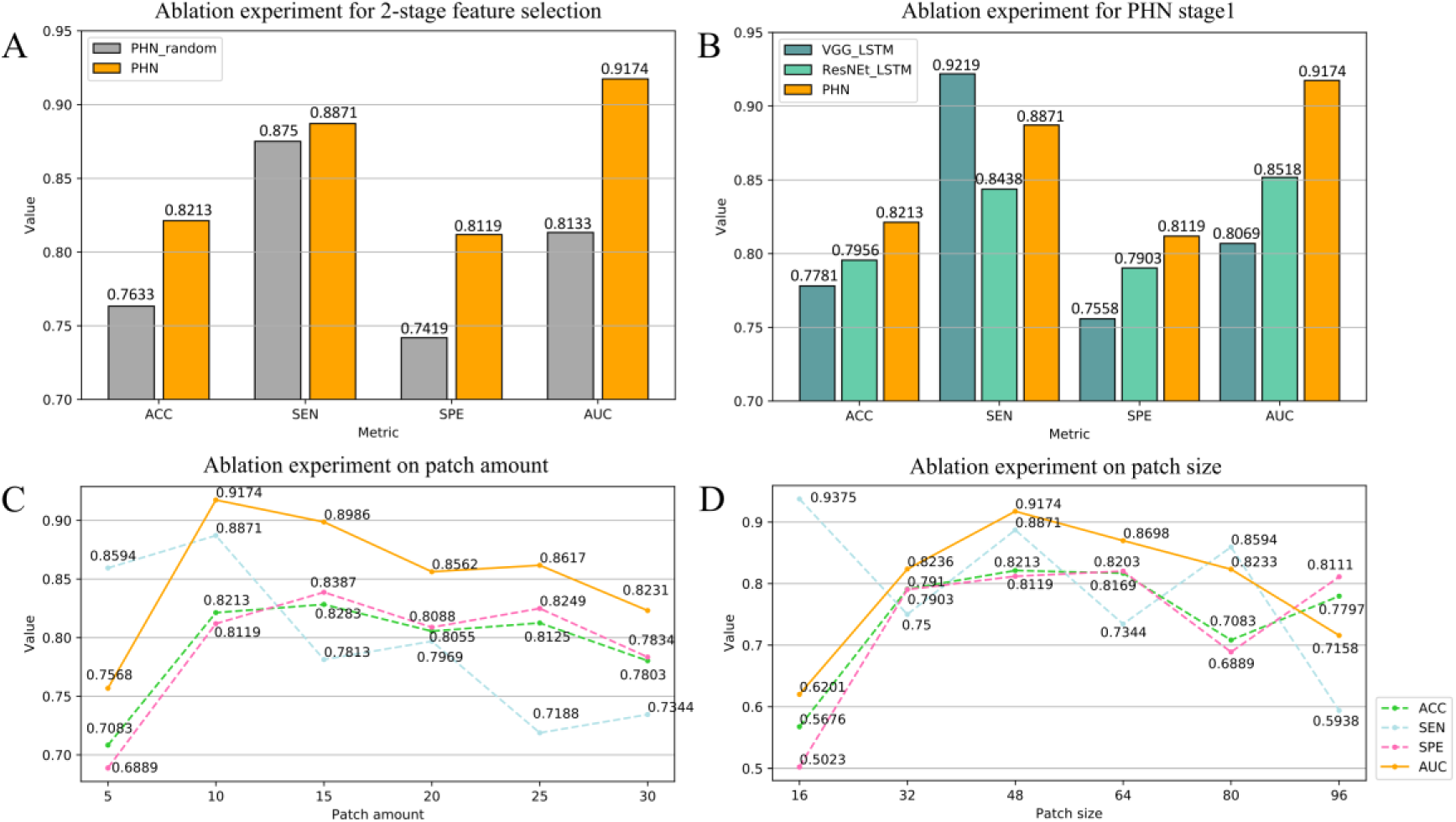
Results of ablation experiments based on the HC-others classification task. (A) The performance difference between randomly taking image blocks in PHN-stage2 (PHN_random) and taking the image blocks according to the confidence order; (B) The performance difference between replacing the P-FEN module with other deep learning network models (PHN_VGG and PHN_ResNet) and the original PHN model; (C) The relationship between performance and the number of input image blocks; (D) The relationship between performance and the size of image blocks. ACC: accuracy; SEN: sensitivity; SPE: specificity; AUC: area under the ROC curve.

We further examined the robustness of PHN to the size (default: 48×48×48) and amount of selected image patches in PHN-stage2 (default: top 10 image patches with the highest classification confidence). Figure 4C reflects that including too few image patches (e.g., top 5) could reduce the performance of PHN. However, when including a sufficient number of image patches, the AUC of PHN remained relatively stable. Figure 4D shows that a range of patch sizes, from 32 to 80, yielded a high AUC of PHN, suggesting the robustness of the algorithm against different patch size parameters.

### Five-category classifications of mental diseases

In the rare cases that require five-alternative choices, PHN could combine the features extracted by the PHN binary classifiers (Figure 1B) to predict the disease label. Each image was input into the 10 PHN binary classifiers between different diagnosis labels and the 5 PHN one vs. others recognition models for feature extraction. The selected features as well as age and gender were concatenated and fed into a 5-layer perceptron to obtain the final 5-class classification results.

Figure 5A shows that the sensitivity metrics for identifying each of the five categories were higher than the random-guess probability (0.12, 0.19, 0.07, 0.52, and 0.11 for HC, BD, MDD, SZ, and OCD, respectively). The overall accuracy was also higher than 50%. The distribution of sensitivity and accuracy was stable among the five-fold cross-validation. The confusion matrix (Figure 5B) indicates that the 5-categroty classifier correctly identified most individuals in each category (the highest proportion in each column is on the diagonal), and the error was due to difficulties in distinguishing specific category pairs, such as OCD-HC, BD-SZ, and MDD-HC. This error pattern suggests the potential to further enhance the performance by combining disease-specific classifiers with different advantages.

**Figure 5.**
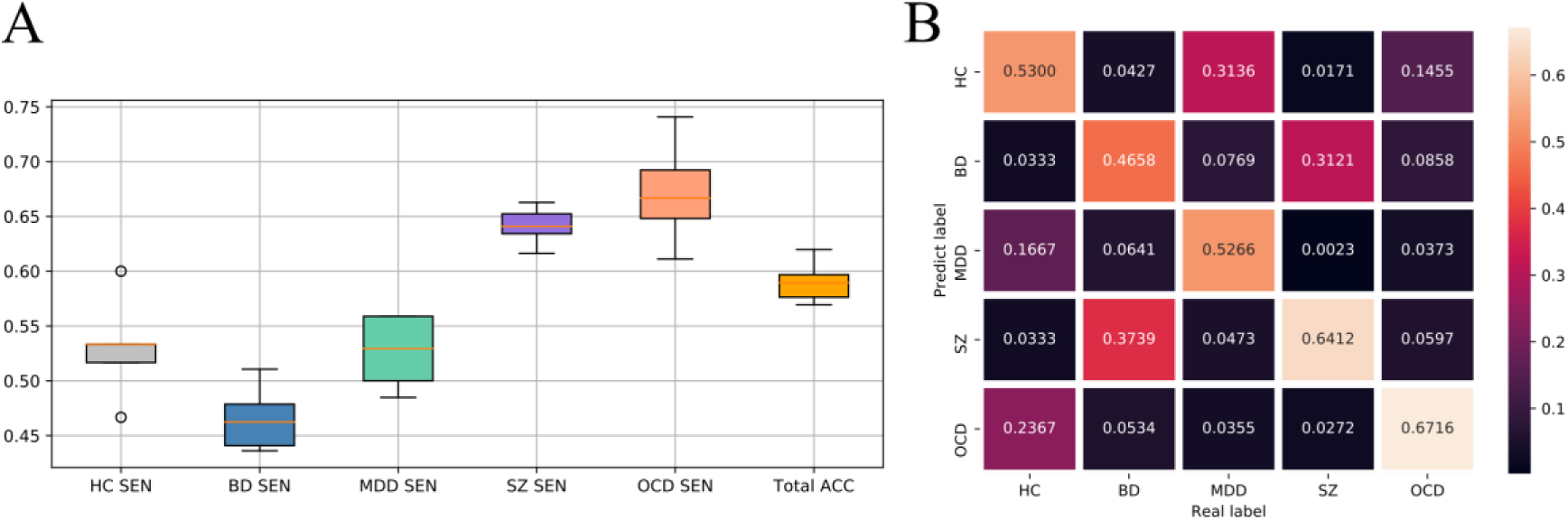
The performance of the 5-category classification model. (A) Sensitivity of each category and total accuracy. The box plot represents the variation in the five-fold cross-validation. (B) Confusion matrix of the classification results. The columns of the matrix represent the actual group labels. Each element in the matrix represents the proportion of correctly identified individuals to the total number of people in the group represented by the column. The proportions of each column add up to 1. ACC: accuracy; SEN: sensitivity.

### Generalizable performance in independent validation

Table 1 shows the performance of PHN classifiers on four open data sets (LA5c^42^, COBRE^43–45^, HBN^46^, and SRPBS^47^) (see Methods). In the LA5c, COBRE, and HBN datasets, the AUCs of BD-HC and SZ-HC classification tasks were above 0.8, and the AUC of MDD-HC and BD-SZ tasks was lower. In the SRPBS dataset, the AUC of the MDD-SZ classification task was 0.69, while the AUC of other tasks was above 0.78. These results support the generalizability of PHN and echo the relative strengths and weaknesses reflected in the cross-validation study.

**Table 1.** Performance of PHN on four open datasets.

| Dataset | Task | ACC | SEN | SPE | AUC |
| --- | --- | --- | --- | --- | --- |
| LA5c<br>(120 HC, 44 SZ, 48 BD) | BD vs HC | 0.7815 | 0.5641 | 0.8875 | 0.8251 |
|  | SZ vs HC | 0.7826 | 0.6 | 0.8625 | 0.8068 |
|  | BD vs SZ | 0.527 | 0.6154 | 0.4286 | 0.5563 |
| COBRE | SZ vs HC | 0.8053 | 0.8364 | 0.7759 | 0.8335 |
| (73 HC, 69 SZ) |  |  |  |  |  |
| **HBN** |  |  |  |  |  |
| (208 HC, 64 MDD, Children) | MDD vs HC | 0.633 | 0.6471 | 0.6287 | 0.6467 |
| **SRPBS** |  |  |  |  |  |
| (926 HC, 38 BD, 237 MDD, 145 SZ) | BD vs HC | 0.8091 | 0.6 | 0.8176 | 0.8217 |
|  | BD vs MDD | 0.7955 | 0.6667 | 0.8158 | 0.7896 |
|  | BD vs SZ | 0.7329 | 0.6333 | 0.7586 | 0.8037 |
|  | MDD vs HC | 0.7882 | 0.7421 | 0.8 | 0.8154 |
|  | MDD vs SZ | 0.6601 | 0.6158 | 0.7328 | 0.6936 |
|  | SZ vs HC | 0.7325 | 0.7672 | 0.727 | 0.7825 |

Abbreviations: LA5c: UCLA Consortium for Neuropsychiatric Phenomics LA5c dataset; COBRE: Centers of Biomedical Research Excellence dataset; HBN: the Healthy Brain Network dataset; SRPBS: the Japanese Strategic Research Program for the Promotion of Brain Science multi-disorder MRI dataset (restricted); ACC: accuracy; SEN: sensitivity; SPE: specificity; AUC: area under the ROC curve.

We also compared PHN classifiers’ performance with those published in previous studies^31–32, 48–55^ (Figure 6). PHN obtained the highest AUC in the SZ vs. HC, OCD vs. HC, and BD vs. HC task in classification performance.

**Figure 6.**
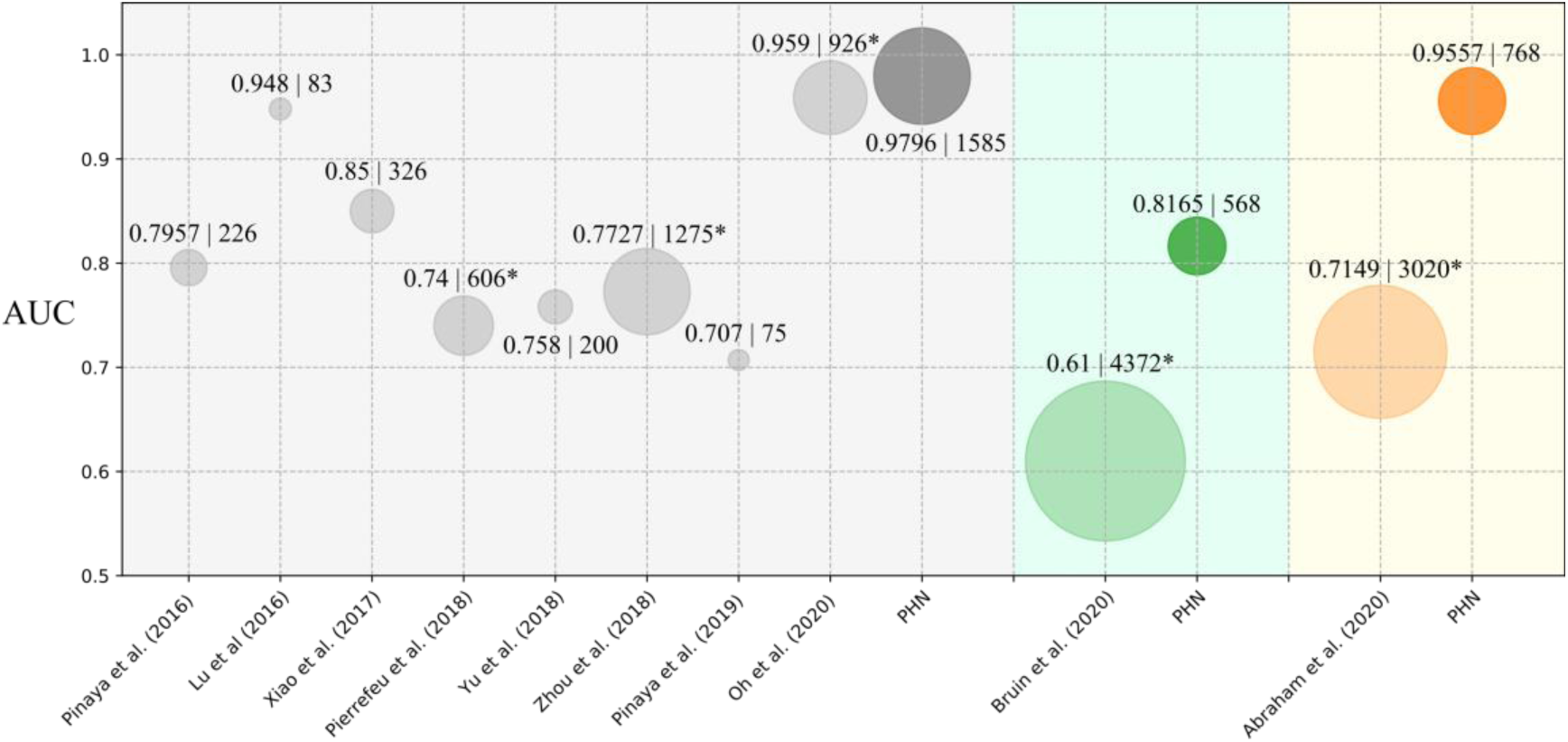
Comparison of the experiments in this paper with some existing work on AUC. Each circle indicates a participating work for comparison, blue indicates SZ vs. HC, orange indicates OCD vs. HC, and green indicates BD vs. HC. The circle’s radius indicates the sample size of the study, and the data label next to the circle is (AUC value | sample size). The * superscript of sample size refers that the study included data from multiple scanning centers. The vertical axis indicates the AUC value obtained for each work.

### Real world implementation and validation

Based on the PHN classifiers, we implemented a functioning application for neuroimaging-aided diagnosis of major mental disorders in Shanghai Mental Health Center. Figure 7A presents a scheme of the system, which automatically connects the physician workstation, MRI workstation, image analysis server. This system achieved a closed loop of psychiatrist’s request, image acquisition, automatic analysis, result delivery, and psychiatrist’s feedback. The PHN classifiers can be further tuned based on the feedback of psychiatrists’ consensus diagnoses. Besides the output of PHN classifiers, this system also conveys the detected brain regions that deviate from the normal range in healthy population in terms of morphological indices (e.g., gray matter volume) and the extent to which these abnormalities are associated with disease diagnoses (odds ratio) (Figure 7B).

**Figure 7.**
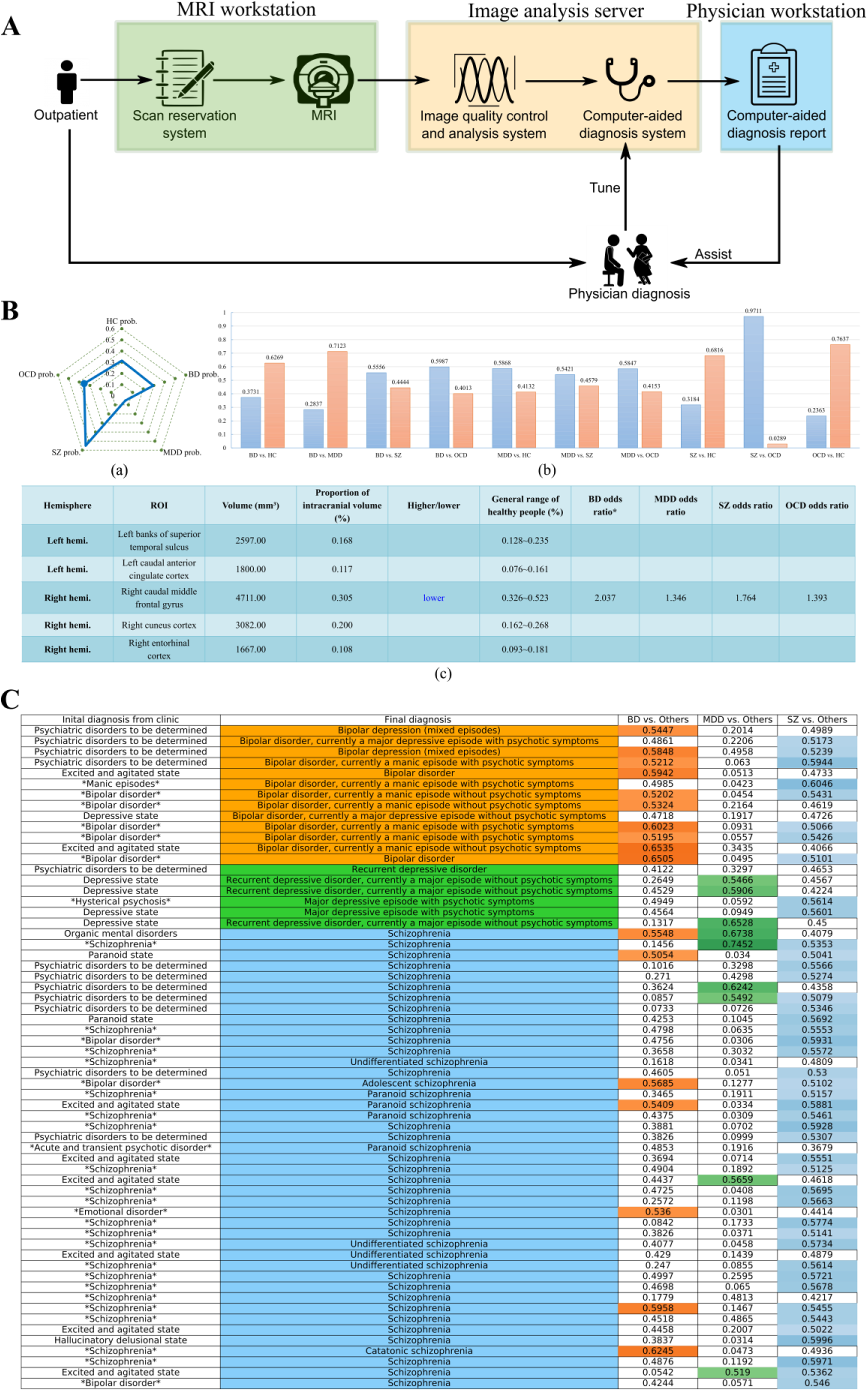
The framework and application of the neuroimaging-aided diagnosis system of major mental disorders in Shanghai Mental Health Center. A: The overall architecture of the system. B: Examples of analysis results that can be acquired by the neuroimaging-aided diagnosis system. (a) Recognition probability distribution of one diagnosis from the others. (b) Transdiagnostic disease prediction probability; (c) Volumetric status, as well as the dominance ratio for various psychiatric disorders of the different brain regions of a certain patient. C: PHN’s performance in a real-world application with 62 patients. The first column presents the initial (transitional) diagnosis from the clinic. The second column presents the consensus diagnosis from a team of psychiatrists based on a series of examinations and observations in the ward. Orange, green, and blue code the diagnostic categories of BD, MDD, and SZ, respectively. The last three columns show the confidence values obtained by the three PHN classifiers. The confidence value> 0.5 is colored, and the transparency of the color encodes the confidence value. *DISEASE TYPE* is the exact diagnosis given by clinicians according to DSM-5 criteria. * Odds ratio characterizes the degree of correlation between the indicator and the current disease. odds ratio>1: the indicator is positively correlated with the occurrence of the current disease; odds ratio<1: the indicator is negatively correlated with the occurrence of the current disease; odds ratio=1: the indicator is not correlated with the occurrence of the current disease.

Based on this clinical application, we validated the performance of PHN in real-world clinical practice. Based on T1-weighted 3D MR brain images of 323 patients (127 MDD, 83 BD, and 113 SZ) collected in the outpatient clinic, the AUC for classification of PHN was 0.6215 for BD-other task, 0.7654 for MDD-other task, and 0.6704 for SZ-other task (Supplementary Figure S2). In addition, we obtained consensus diagnoses for 62 of 323 patients (6 MDD, 13 BD, and 43 SZ), which were made by a team of psychiatrists based on a comprehensive examination and observations on the ward. Based on the consensus diagnosis and decision cutoff = 0.5, the classification accuracy of PHN was 0.8387 for BD-other tasks, 0.8548 for MDD-other tasks, and 0.6935 for SZ-other tasks. As shown in Figure 7C, a large portion of the errors made by the PHN classifier were accompanied by co-morbidities in the consensus diagnoses, such as “bipolar disorder with psychotic symptoms”. The output of the PHN classifier echoes this co-morbidity phenomenon, suggesting that the classifier correctly reflects the complexity of psychiatric diagnoses in the real world.

## Discussion

The complex symptoms of mental disorders and the lack of objective diagnostic tools make psychiatric diagnosis difficult. Here, we demonstrate for the first time that the combination of structural brain images with deep learning network models can help solve the problem of diagnosing multiple major psychiatric disorders in the real world. We propose a deep learning network, PHN, and apply it to a large-scale cross-disease brain imaging dataset to train a series of classifiers that can identify any of the classes of HC, MDD, BD, SZ, and OCD. These classifiers showed good performance on cross-validation, independent public data, and real-world clinical data. This study achieves the ability to accurately identify one of multiple diagnoses and allows multiple diagnoses to coexist, thus addressing a real-world need in psychiatric practice. Based on these efforts, we have implemented a closed-loop system for neuroimaging-assisted diagnosis of major psychiatric disorders and have initially validated its effectiveness in real-world applications. Thus, this research contributes to the translation of brain research into an objective diagnostic aid for psychiatric disorders.

### Identify specific diagnoses among multiple mental disorders and allow for comorbidities

A novel feature of the PHN adapted to the needs of clinical applications is its ability to accurately identify one type of diagnosis from among multiple types of psychiatric disorders (Figure 2B). This feature is better suited to address the practical clinical needs of psychiatrists who need to weigh multiple diagnoses than the results of most previous studies that focused on two groups. Meanwhile, the PHN classifiers enable parallel inference of different diagnostic categories, thus allowing more than one diagnostic inference for the same individual. This feature also makes the output of the PHN classifier more consistent with the highly co-morbid clinical characteristics of mental disorders^4, 6^. Besides, PHN classifiers also perform well on most 2-category classification tasks, so psychiatrists can only refer to the PHN classifiers’ recommendations for competing diagnostic types.

### PHN’s design and the cross-disease training dataset help improve classifier performance

Most of the machine learning studies on brain structure imaging of SZ, MDD, BD, and OCD combined conventional classifiers (such as support vector machine, SVM) and hand-crafted image features (e.g., voxel-based morphometrics, regional gray matter volume, and image features based on spatial deformation, tensor analysis, and shape analysis) ^29–31, 33–38^. Research in other fields^56–58^ has shown that deep learning is more accurate than traditional neuroimaging classification algorithms. However, the few current studies applying deep learning methods have used hand-crafted image features^39^ or skipped active feature selection -- treating the whole-brain image data equally^32^. Hand-crafted features may overwhelm fine-grained structural features of individuals due to spatial manipulation and averaging over regions. Accordingly, some studies have supported the advantages of voxel-level analysis over region-based analysis^59–60^. Yet, given the limited sample size in the psychiatric imaging field, equally treating the voxels of the whole brain may cause information redundancy and reduce training effectiveness. Therefore, deep learning methods with attention mechanisms are expected to help improve the performance of classifiers.

PHN is tailored to the specific needs of neuroimaging classification for mental disorders, resulting in PHN classifiers outperforming several other traditional machine learning and deep learning methods for all classification tasks (Figure 3). First, voxel-level features may represent physiological information and play essential roles in image feature-based classifiers^59–60^, but aligning images to a standard space or averaging signals within regions may disturb voxel-level features. PHN evaluates and selects local features (spatial patches) by training a deep learning model (P-FEN), thus avoiding the voxel-level interpolation and averaging process. Figure 4B provides empirical evidence for the effectiveness of P-FEN, and Figure 4C-D support that the performance of PHN is stable for the number and size of spatial patches, unless they are too small. Second, with limited sample size, equally applying whole-brain information to train classifiers may lead to redundancy and reduce the training efficiency^62–63^. The attention stage of PHN helps to reduce the information redundancy in classifier training. This advantage is supported by comparing two deep learning algorithms (VGG^64^ and ResNet^65^) that process the whole brain image uniformly (Figure 3A-B). Figure 4A also shows the essential role of pre-selection and ranking of spatial patches in PHN-stage1. Third, the brain works as an integrated system, and local brain abnormalities are interrelated^98–101^. Thus, treating spatially distributed abnormalities as independent predictors may lose essential information. PHN’s new application of the LSTM module solves this issue by treating the interdependence of spatially separated patches as a sequence. The results in Supplementary Table S2 show that the LSTM module in PHN-stage 2 accommodates an overall better performance than traditional fully connected neural networks.

In addition, training on a carefully labeled large cross-disease dataset obtained on the same scanner helps to improve the effectiveness of the PHN. The dataset we used is a large-scale dataset covering a wide range of mental disorders and contains data from carefully labeled and quality-controlled studies. The data were enrolled by 10 psychiatric study groups according to strict criteria, using essentially the same scanning process and parameters, undergoing fine quality control, and processed and managed using the same hardware and software platform^66^. Data from the same location has advantages in terms of image quality and consistency in socio-cultural context and understanding of diagnostic criteria, which helps to reduce noise in identifying specific diseases. Applying the PHN to data from multiple diagnoses also makes it easier for the PHN to identify features of specific diagnostic categories. The advantages of classifier design and training dataset facilitate the performance of the PHN classifiers, not only in cross-validation and independent validations, but also beyond previous studies^31–32, 48–55^ (Figure 6).

### The performance of PHN classifiers is generalizable and effective in real world applications

The cross-site generalization of the PHN classifier is supported by the validation using four independent datasets (Table 2). This result shows that with some fine-tuning, the PHN classifier can be applied to neuroimaging data collected using different scanners. This feature is important because site effects have a significant impact on the systematic bias of image segmentation, thus hurting the cross-site generalizability of neuroimaging markers based on image segmentation^67^. Although researchers have been working to harmonize site effects in image segmentation, there is no consensus on an effective solution^68–72^. PHN classifiers do not require an explicit image segmentation step, but rather embed possible site effects in local features through deep learning. It has been suggested that machine learning methods may be effective in tuning site differences, and our empirical evidence supports the cross-site generality of PHN classifiers. Since the sample size and diagnosis type of independent open datasets limited the amount of data used to tune the PHN, we expect that the cross-site performance of the PHN can be further improved with larger samples.

**Table 2.**
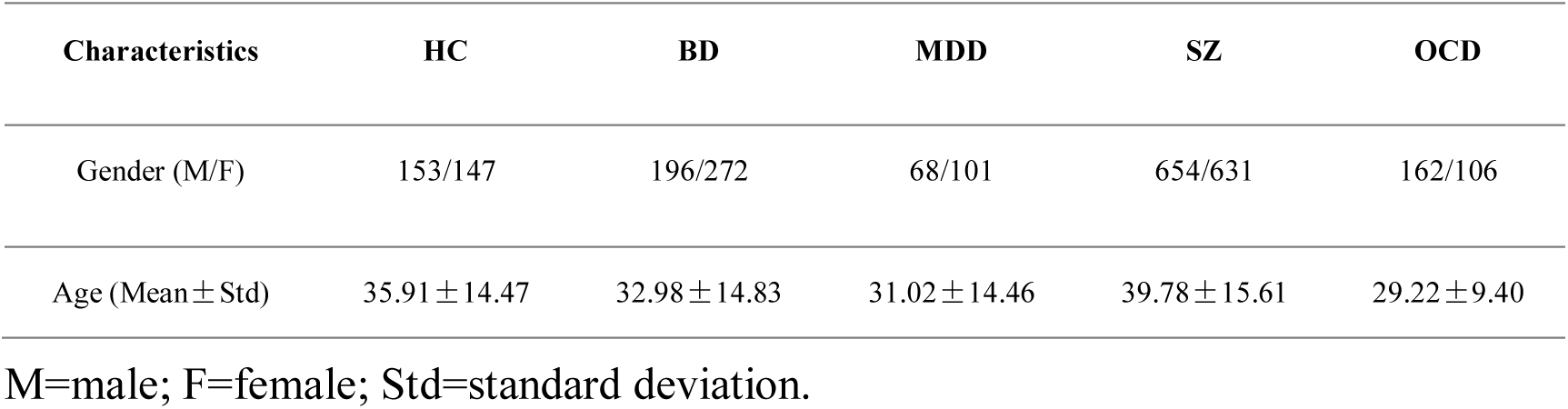
The demographic characteristics of 2490 subjects.

Importantly, the real-world validation shed light on the application of PHN to real-world clinical problems that are much more complex than those in case-control group comparison studies. Subject to many factors, the initial diagnosis in clinical practice is often symptom suggestive and different from the final consensus diagnosis. Furthermore, it is common for symptoms to overlap (e.g., a patient diagnosed with BD or MDD may also have concurrent psychotic symptoms, Figure 7C). These facts may increase the inconsistency between diagnoses made by different psychiatrists. PHN provides an objective measure and evidence-based reference for diagnosis. Although classification performance is lower in real-world data than in rigorously cleaned scientific data (especially when the initial diagnosis is used as the diagnostic label), in most cases, PHN results are consistent with the consensus diagnosis. For those patients with overlapping symptoms, the discriminant probabilities for each disease category from the PHN reflect co-morbidities accordingly. Thus, going beyond case-control studies based on clean diagnostic labels, a novel contribution of this study is to achieve the translation from neuroimaging biomarker research to clinical practice with acceptable accuracy.

In addition, the consistency of PHN with the initial diagnosis was verified on 323 real-world patients. As seen in Supplementary Figure S2, while the AUC greater than 0.5 was achieved in the identification tasks of BD, MDD, and SZ, the diagnostic performance of PHN was unsatisfactory when the initial diagnosis was considered as the gold standard. An important reason for these results is the possible confusion between diseases in the initial diagnosis. Another piece of evidence supporting this inference is that the ROC curve reveals that most patients with an initial diagnosis labeled MDD tend to be predicted by the PHN to be in the “other category”: in practice, the initial diagnosis tends to be conservative (a patient will not be classified as having a serious mental illness such as BD or SZ at the outpatient clinic unless the specific symptoms are extremely obvious), so “MDD patients” used to test for PHN were actually likely to be in another mental illness category.

### A closed-loop system of neuroimage-assistant diagnosis system for major mental disorders

With PHN as the core engine, we have implemented a closed-loop neuroimaging-assisted diagnostic system. We have integrated it in a clinical workflow to validate and improve the performance and stability of the system in a real-world setting (Figure 7). To our best knowledge, this is the first cross-disease neuroimaging-assisted diagnostic system translated into a clinical service test. This system not only provides objective diagnostic reference for psychiatrists, but also enables continuous learning from physician feedback and patient progression records to improve accuracy and upgrade predictive capabilities. Based on the accumulation of large amounts of data, the system also suggests individual reports of abnormalities at the brain region level and the strength of their association with disease diagnosis (Figure7A-B), thus providing psychiatrists with more intuitive and interpretable objective cues. We expect this neuroimaging-assisted diagnostic system to provide a truly usable and objective artificial intelligence reference for psychiatric clinical services as it is continuously improved and validated.

### Future directions

Our primary considerations for using high-resolution T1w brain structure images as a classification feature: 1) this imaging modality shows high reliability^73^; 2) mental disorders show abnormalities in this imaging modality^31–32, 48–55^; and 3) imaging techniques are already available to help reduce their scan time and enable assessment of head movement levels during the scan, thus increasing the likelihood of successful acquisition of high-quality images. Future work could involve abnormalities in brain function as additional diagnostic features besides other brain structural modalities. Some functional brain imaging paradigms, such as naturalistic imaging^74^, have been shown to have high reliability and are relatively easy to implement, which may help to evoke abnormalities in brain working patterns.

This work uses current psychiatric diagnostic systems as the “gold standard” for implementing an artificial intelligence system that could assist current clinical practice. However, there are also nosologies and theories in psychiatry that differ from clinical diagnostic systems, such as HiTOP^75^ and RDoC^76^, to address the high heterogeneity and co-morbidity in the current diagnostic system. Future research could explore neuroimaging systems that detect different symptom dimensions and infer the benefits of other treatment options, thus making neuroimaging-assisted diagnostic systems more functional.

In addition, the classification performance between some diagnostic categories in this work needs further improvement, such as the MDD vs. HC and BD vs. SZ tasks. These phenomena are consistent with previous findings^77–78^, and thus they may suggest pathological ambiguity. Future studies could focus on improvements in terms of underlying theory and technology.

In conclusion, this study has developed and verified a neuroimaging model fitting the clinical needs of psychiatric diagnosis. Based on this model, we build a closed-loop neuroimaging-assistant diagnostic system embedded in the clinical process. The effectiveness and applicability of this system has been verified in the real world application.

## Methods

### SMHC dataset

T1-weighted high-resolution brain structural images of healthy controls (HC) and patients with BD, MDD, SZ, OCD diagnoses were collected in Shanghai Mental Health Center (SMHC) in China, using a Siemens 3.0 Tesla Verio scanner with highly similar acquisition parameters (Supplementary Table S3). The participants were initially recruited by ten research groups of SMHC from local clinics/communities for various research projects from April 2015 to May 2020. All participants (or legal guardians) signed written informed consent during the recruitment. The IRB committee of Shanghai Mental Health Center approved this study and permitted the use of measures extracted from the coded data (IRB2020-15). The following inclusion criteria for the participants were applied: (1) 18-59 years old; (2) A diagnosis of SZ, OCD, BD, or MDD according to Diagnostic and statistical manual of mental disorders (DSM-5) ^79^, DSM-IV, and the International Classification of Diseases (ICD)-10^80^. A diagnosis of BD respectively based on the DSM-5 (n =271), DSM-IV (n = 61), and the ICD-10 (n = 136); The patients were diagnosed with MDD using the DSM-IV (n = 169); The patients were diagnosed with SZ using the DSM-5 (n = 39), DSM-IV (n = 95), and the ICD-10 (n = 1151); The patients were diagnosed with OCD using the DSM-5 (n = 205), DSM-IV (n =63); (3) Completed a high-resolution T1-3D brain scan that passed the quality check.

The exclusion criteria were: (1) any current or lifetime Axis I psychiatric disorder comorbidity screened by Mini-International Neuropsychiatric Interview (MINI)^81^; (2) a history of substance abuse and serious physical disease. The healthy controls were recruited from local communities. They underwent a MINI interview to exclude any mental disorder and had no history of antipsychotic medication and serious physical diseases.

Senior radiologists checked all images included in this study to exclude obvious brain structural abnormalities and imaging artifacts. The quality of the images was further quantified using CAT12.6 toolbox^82^ based on the resolution, intensity bias, and noise level. Images with a quality grade lower than ‘B-’ were excluded from further analyses. After quality control, a total of 2490 data of participants, including 300 HC subjects, 468 BD patients, 169 MDD patients, 1285 SZ patients, and 268 OCD patients, were included in the subsequent analyses (Table 2).

The images were resampled into a resolution of 256×256×256 and corrected for intensity inhomogeneity using the N3 algorithm^83^. Then we performed skull-stripping using watershed algorithm^84^ and a process of manually checking to ensure that the skull was completely removed. All the above processing steps were performed using the same hardware and software platform^66^.

### Open research datasets

We used four open datasets to examine the generalizability of PHN’s performance. The datasets included: 1) the UCLA Consortium for Neuropsychiatric Phenomics LA5c (LA5c) dataset^42^, which contains 120 HC subjects, 44 SZ subjects, and 48 BD subjects; 2) Centers of Biomedical Research Excellence (COBRE) dataset^43–45^, which contains 73 HC and 69 SZ subjects, 3) the Healthy Brain Network (HBN) dataset (release 1-9)^46^, which contains 64 MDD and 208 HC children and teenagers, and 4) the Japanese Strategic Research Program for the Promotion of Brain Science (SRPBS) multi-disorder MRI dataset (restricted)^47^, which contains 926 HC subjects, 38 BD subjects, 237 MDD subjects, and 145 SZ subjects. Details of these datasets are presented in Supplementary Table S4, and the quality control process and preprocessing steps are exactly the same as the process described above. Due to the lack of an open neuroimaging dataset for OCD, the OCD-related performance was not examined. The quality control process and preprocessing steps were fully consistent with the process described earlier. To reduce the influence of scanner differences on the results, we randomly used 20% of each public dataset to pre-train (fine-tune) the parameters of PHN, and the rest of the data were used as test data.

### Real-world validation data

The greatest value of machine learning models lies in the integration of high-throughput information for practical services, but most neuroimaging-based classification models of mental illness, including PHN, were trained and verified based on data collected in accordance with scientific research standards. Since most research standards exclude comorbidities, ambiguous diagnoses, etc., it is not sufficiently representative of the real-world clinical situation.

We verified the performance of PHN in real-world clinical practice. We collected T1-weighted 3D MR brain images of 323 patients (83 BD, 127 MDD and 113 SZ, Supplementary Table S5), and their (interim) diagnosis obtained in the outpatient clinic. In addition, 62 patients (6 MDD, 13 BD, and 43 SZ) among 323 patients have consensus diagnosis (according to DSM-5 criteria) made by a team of psychiatrists based on comprehensive examinations and observations in the ward. The quality control process and preprocessing steps are the same as the process described above. PHN predicts the diagnosis based on brain images and evaluates real-world performance based on the interim and final diagnosis in the ward.

### Patch-based hierarchical network (PHN)

As demonstrated in Figure 1A, we developed a deep learning framework, patch-based hierarchical network (PHN), to recognize the images’ diagnosis labels. PHN contains two stages: PHN-stage1 detects diagnosis-sensitive brain locations, and PHN-stage2 performs local feature extraction and classification. The rationale behind this design is that pre-locating possible disease-related brain regions (PHN-stage1) may help reduce redundant information and improve classification performance (PHN-stage2).

PHN-stage1 randomly extracts 300 image patches with a size of 48×48×48 from the input image and feeds them into the trained P-FEN model to obtain disease probabilities. The centers of the patches are constrained within the brain. Based on the predicted probabilities of the 300 patches, an individual-specific confidence map (i.e., the attention map) is generated. Specifically, the diagnosis confidence with respect to the *i*-th voxel in the brain image has the following value *C*_*i*_:

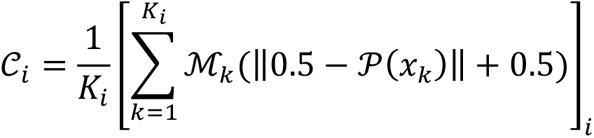

where *K*_*i*_ denotes the frequency of the *i*-th voxel in the 300 patches, ℳ_*k*_ (*t*) : [0.5, 1] → ℝ^256×256×256^ refers to the operator that adds the value of *t* ∈ [0.5, 1] to the patch with an appropriate location in a zero-padded image in ℝ^256×256×256^, and 𝒫(*x*_*k*_) ∈ [0, 1] denotes the disease prediction probability for the *k*-th image patch. Here, ‖0.5 − 𝒫(*x*_*k*_)‖ calculates the distance between 0.5 and 𝒫(*x*_*k*_), as the prediction probability of 0.5 refers to the random guess while the probabilities close to 0 and 1 indicate the prediction with high confidence; 0.5 was further added to ensure the calculated confidence value ranges is between 0.5 to 1, which 0.5 indicates the absence of any categorical tendency, while 1 indicates a strong conviction of categorical tendency. Supplementary Figure S1 presents examples of such voxel-wise attention maps.

### Model training and cross-validation

The PHN model was implemented using Python 3.7 based on PyTorch 1.6.0, and the computer we used contains a single GPU (i.e., NVIDIA Quadro P5000 16 GB), and the operating system is CentOS 7.9. Classifiers that distinguish every two groups among HC, BD, MDD, SZ, and OCD groups, as well as one group from the others, were trained separately. The classifiers were trained and tested using a 5-fold cross-validation scheme. The samples in each diagnosis group were divided into 80% of the training set and 20% of the testing set in each fold. Within the training set, 2/3 of the samples (keeping the ratio of the two groups) were used to train the P-FEN model for 300 epochs, and the other 1/3 samples were used to evaluate the performance and select the best model. The trained P-FEN was then deployed to PHN-stage2, and the same procedure and data as above were used to train PHN-stage2. For all training steps, the adaptive moment estimation algorithm (ADAM)^85^ was used as the optimizer with a learning rate of 1 × 10^−4^, and the batch size was set to 28.

### Performance metrics

We computed sensitivity (SEN), specificity (SPE), the area under the receiver operating characteristic curve (AUC), F1-score (F1), and Matthews correlation coefficient (MCC) in each test set in the cross-validation. The F1-score and MCC are defined as:

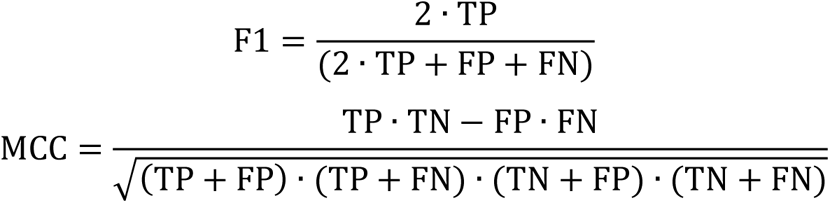

where TP and TN denote true positive and true negative values, and FP and FN refer to false positive and false negative values, respectively. The F1 and MCC are both measures for the classification performance of the 2-class classification task. F1 considers both precision and recall, and MCC serves as a balanced measure of classes of different sample sizes. The mean and standard deviation values of these metrics were calculated among the five folds of cross-validation.

### Comparisons with other methods

We compared the performance of PHN with five widely-used machine learning methods, including 1) logistic regression, with L2 penalty, C=1, and 1e-4 of tolerance for stopping criteria; 2) support vector machine (SVM)^86^, with linear kernel, C=1, and 1e-3 of stopping tolerance; 3) random forest classifier^87^, with 100 trees, Gini impurity as split criteria, 2 for the minimum number of samples required to split an internal node, and 1 for the minimum number of samples required to be at a leaf node; 4) extreme gradient boosting (XGBoost)^88^, with maximum tree depth=2, learning rate=0.1, estimator number=100; 5) gradient boosting decision tree (GBDT)^89^, with deviance as loss, 0.1 as learning rate, estimator number=100, Friedman-MSE as a criterion.

To derive imaging features for these methods, we used FreeSurfer (version 6.0.0)^90^ to compute volume, cortical thickness (for cortical structures only), and cortical surface area (for cortical structures only) measures for brain regions (87 regions) defined in the Desikan-Killiany atlas^91^+subcortical atlas^92^. In addition, we extracted 3D image histological features for the 87 brain regions by using Radiomics (version 3.0.1)^93^. To avoid overfitting, the two sets of features were separately reduced to eight dimensions by principal component analyses and combined into a 16-dimensional feature set. The machine learning models above were then trained using this feature set.

We further compared PHN with two other deep learning models: a 3D visual geometry group network (VGG)^64^, which has been widely used in brain disease diagnosis^94–97^, and a 3D residual network (ResNet)^65^, which has been proved to be the most effective convolutional neural network in computer vision tasks. We slightly modified the structure of VGG and ResNet (see Supplementary Table S6-7) to make them similar to the networks used in existing studies^23–25^ and to reduce the computational cost.

### Ablation studies on the framework structure

To examine the effectiveness of PHN, we performed a series of ablation studies on the network structure. First, to verify the effectiveness of the two-stage framework (i.e., the first stage for brain region localization and the second stage for feature extraction and classification), we trained another 2-stage PHN model for the HC vs. Others task. In this model, ten image patches in PHN-stage2 were randomly selected (rather than selected based on prediction confidence) for further feature extraction. Second, to examine the effectiveness of the P-FEN structure, we replaced the P-FEN with the aforementioned VGG and ResNet in PHN. Third, since we specified the size of the image patches (48×48×48) for P-FEN and the number of image patches (top 10) in PHN-stage2, we conducted a series of experiments to examine the robustness of PHN to the size and amount of the image patches. Fourth, in order to verify the effectiveness of the LSTM model in PHN-stage2, we replaced the LSTM model with a multi-layer perceptron. Finally, we examined the effect of including age and sex information by removing the age and sex input in the final perceptron in PHN-stage2.

### Multi-class classification

We further evaluated the capability of PHN to solve the five-class problem by combining the features extracted by different PHN classifiers (Figure 1B). Each image was input into the ten aforementioned PHN binary classifiers between different diagnosis labels and the 5 PHN recognition model for each disease label (i.e., the current label vs. others). Each of the 15 PHN models extracted a 160-dimensional feature set, and these sets, as well as age and gender information, were concatenated. Then, the concatenated features (dimension is 160×15+2=2402) were fed into a 5-layer perceptron to obtain the final 5-class classification results.

## Data Availability

All data produced in the present study are available upon reasonable request to the authors

## Notes

### Competing Interest Statement

The authors have declared no competing interest.

### Funding Statement

This work was supported by the National Key R&D Program of China (2018YFC2001605); National Natural Science Foundation of China (81971682, 81571756); Natural Science Foundation of Shanghai (20ZR1472800); Shanghai Municipal Commission of Education-Gaofeng Clinical Medicine Grant Support (20171929); Shanghai Clinical Research Center for Mental Health (19MC1911100); Shanghai Municipal Health Commission (2019ZB0201), Shanghai Science and Technology Commission (18JC1420305), Hundred-Talent Fund from Shanghai Municipal Commission of Health (2018BR17); Shanghai Mental Health Center Clinical Research Center (CRC2018DSJ01-5; CRC2019ZD04); Research Funds from Shanghai Mental Health Center (13dz2260500, 2018-YJ-02); 2021 Shanghai Mental Health Center Hospital Level Key Projects（2021zd02).

### Author Declarations

IRB, Shanghai Mental Health Center gave ethical approval for this work.

### Summary of Updates

Adding validation results on 323 real-world data

