## Supplementary Figure S1 for "Deep learning system for brain image-aided diagnosis of multiple major mental disorders"

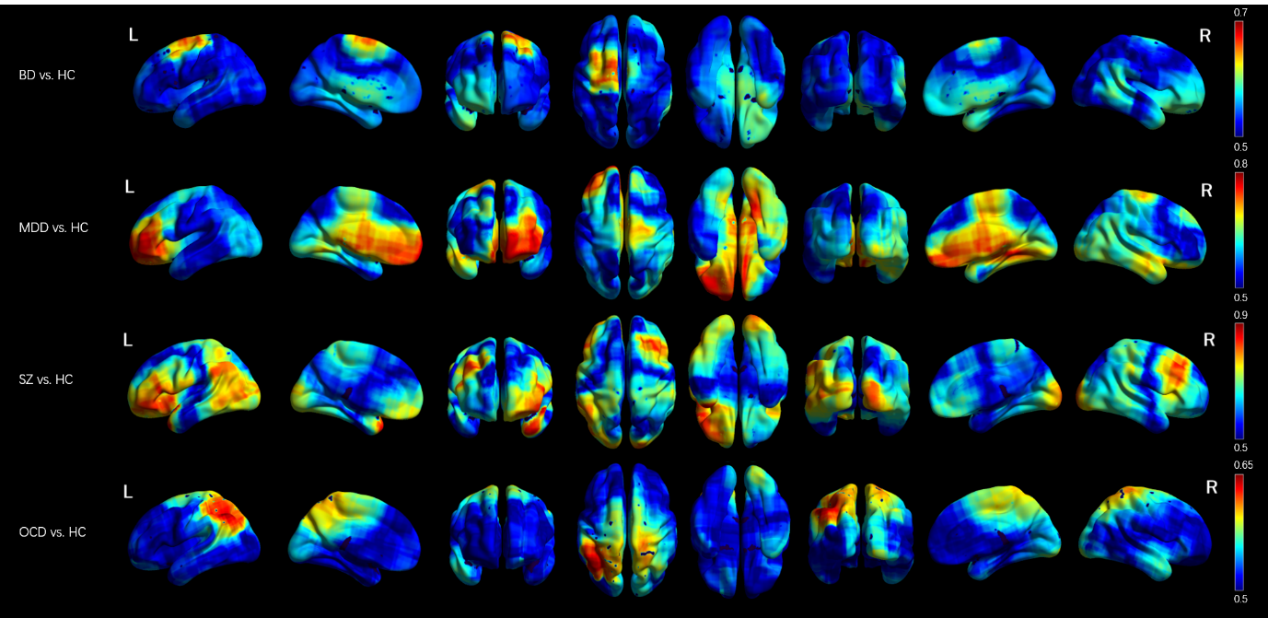

**Supplementary Figure S1** Attentional maps generated by four subjects from the BD, MDD, SZ, and OCD populations in each of the four classification tasks.
