## Supplementary figures and images for "Deep learning system for brain image-aided diagnosis of multiple major mental disorders"

### Supplementary Figure S2

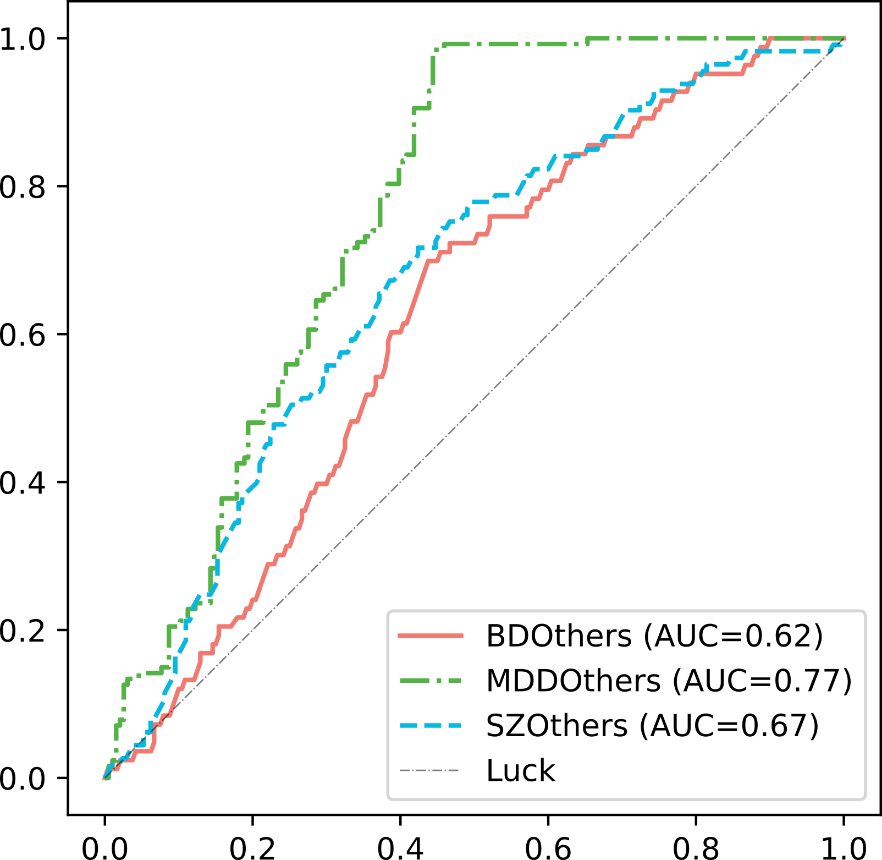

**Supplementary Figure S2** PHN’s performance in a real-world application with 323 patients.
