## Supplementary Table S1 for "Deep learning system for brain image-aided diagnosis of multiple major mental disorders"

**Supplementary Table S1** Details of the P-FEN.

Note: kernel_size is set as 3×3×3 for all Convolution layers; eps is set as 1e-5 and momentum is set as 0.1 for all BatchNorm3D layers.

P_FEN

| Layer | Input | Output | Input channel | Output channel |
| --- | --- | --- | --- | --- |
| RSU7 | x | Hx1 | 1 | 8 |
| Maxpool1 | Hx1 | Hx1_out | 8 | 8 |
| RSU6 | Hx1_out | Hx2 | 8 | 32 |
| Maxpool2 | Hx2 | Hx2_out | 32 | 32 |
| RSU5 | Hx2_out | Hx3 | 32 | 64 |
| Maxpool3 | Hx3 | Hx3_out | 64 | 64 |
| RSU4 | Hx3_out | Hx4 | 64 | 128 |
| Maxpool4 | Hx4 | Hx4_out | 128 | 128 |
| RSU5 | Hx4_out | Hx5 | 128 | 256 |
| AdaptiveMaxpool | Hx5 | Hx5_out | 256 | 256 |
| linear | Hx5_out | Linear_out | 256 | 2 |
| softmax | Linear_out | out | 2 | 2 |
| Return | - | out | - | 2 |

*RSU: residual U-block

*Kernel size and stride of all maxpooling layers are 2.

RSU7

| Layer | Input | Output | Input channel | Output channel |
| --- | --- | --- | --- | --- |
| REBNCONV1 | x | Hx1 | 1 | 8 |
| Maxpool1 | Hx1 | Hx1_out | 8 | 8 |
| REBNCONV2 | Hx1_out | Hx2 | 8 | 8 |
| Maxpool2 | Hx2 | Hx2_out | 8 | 8 |
| REBNCONV3 | Hx2_out | Hx3 | 8 | 8 |
| Maxpool3 | Hx3 | Hx3_out | 8 | 8 |
| REBNCONV4 | Hx3_out | Hx4 | 8 | 8 |
| Maxpool4 | Hx4 | Hx4_out | 8 | 8 |
| REBNCONV5 | Hx4_out | Hx5 | 8 | 8 |
| REBNCONV6 | Hx5 | Hx6 | 8 | 8 |
| REBNCONV5_d | Concatenate(Hx5, Hx6) | Hx5_d | 8+8 | 8 |
| Upsample5 | Hx5_d | Hx5_d_up | 8 | 8 |
| REBNCONV4_d | Concatenate(Hx5_d_up, Hx4) | Hx4_d | 8+8 | 8 |
| Upsample4 | Hx4_d | Hx4_d_up | 8 | 8 |
| REBNCONV3_d | Concatenate(Hx4_d_up, Hx3) | Hx3_d | 8+8 | 8 |
| Upsample3 | Hx3_d | Hx3_d_up | 8 | 8 |
| REBNCONV2_d | Concatenate(Hx3_d_up, Hx2) | Hx2_d | 8+8 | 8 |
| Upsample2 | Hx2_d | Hx2_d_up | 8 | 8 |
| REBNCONV1_d | Concatenate(Hx2_d_up, Hx1) | Hx1_d | 8+8 | 8 |
| Return | - | Hx1_d+Hx1 | - | 8 |

*REBNCONV: ReLU-batch normalization-convolution block

*Kernel size and stride of all maxpooling layers are 2.

RSU6

| Layer | Input | Output | Input channel | Output channel |
| --- | --- | --- | --- | --- |
| REBNCONV1 | x | Hx1 | 8 | 8 |
| Maxpool1 | Hx1 | Hx1_out | 8 | 8 |
| REBNCONV2 | Hx1_out | Hx2 | 8 | 8 |
| Maxpool2 | Hx2 | Hx2_out | 8 | 8 |
| REBNCONV3 | Hx2_out | Hx3 | 8 | 8 |
| Maxpool3 | Hx3 | Hx3_out | 8 | 8 |
| REBNCONV4 | Hx3_out | Hx4 | 8 | 8 |
| REBNCONV5 | Hx4 | Hx5 | 8 | 8 |
| REBNCONV4_d | Concatenate(Hx5, Hx4) | Hx4_d | 8+8 | 8 |
| Upsample4 | Hx4_d | Hx4_d_up | 8 | 8 |
| REBNCONV3_d | Concatenate(Hx4_d_up, Hx3) | Hx3_d | 8+8 | 8 |
| Upsample3 | Hx3_d | Hx3_d_up | 8 | 8 |
| REBNCONV2_d | Concatenate(Hx3_d_up, Hx2) | Hx2_d | 8+8 | 8 |
| Upsample2 | Hx2_d | Hx2_d_up | 8 | 8 |
| REBNCONV1_d | Concatenate(Hx2_d_up, Hx1) | Hx1_d | 8+8 | 32 |
| REBNCONV_in | x | Hx1_in | 8 | 32 |
| Return | - | Hx1_d+Hx1_in | - | 32 |

RSU5

| Layer | Input | Output | Input channel | Output channel |
| --- | --- | --- | --- | --- |
| REBNCONV1 | x | Hx1 | 32 | 16 |
| Maxpool1 | Hx1 | Hx1_out | 16 | 16 |
| REBNCONV2 | Hx1_out | Hx2 | 16 | 16 |
| Maxpool2 | Hx2 | Hx2_out | 16 | 16 |
| REBNCONV3 | Hx2_out | Hx3 | 16 | 16 |
| REBNCONV4 | Hx3 | Hx4 | 16 | 16 |
| REBNCONV3_d | Concatenate(Hx4, Hx3) | Hx3_d | 16+16 | 16 |
| Upsample3 | Hx3_d | Hx3_d_up | 16 | 16 |
| REBNCONV2_d | Concatenate(Hx3_d_up, Hx2) | Hx2_d | 16+16 | 16 |
| Upsample2 | Hx2_d | Hx2_d_up | 16 | 16 |
| REBNCONV1_d | Concatenate(Hx2_d_up, Hx1) | Hx1_d | 16+16 | 64 |
| REBNCONV_in | x | Hx1_in | 32 | 64 |
| Return | - | Hx1_d+Hx1_in | - | 64 |

RSU4

| Layer | Input | Output | Input channel | Output channel |
| --- | --- | --- | --- | --- |
| REBNCONV1 | x | Hx1 | 64 | 32 |
| Maxpool1 | Hx1 | Hx1_out | 32 | 32 |
| REBNCONV2 | Hx1_out | Hx2 | 32 | 32 |
| REBNCONV3 | Hx2_out | Hx3 | 32 | 32 |
| REBNCONV2 | Concatenate(Hx3, Hx2) | Hx2_d | 32+32 | 32 |
| Upsample2 | Hx2_d | Hx2_d_up | 32 | 32 |
| REBNCONV1_d | Concatenate(Hx2_d_up, Hx1) | Hx1_d | 32+32 | 128 |
| REBNCONV_in | x | Hx1_in | 64 | 128 |
| Return | - | Hx1_d+Hx1_in | - | 128 |

RSU5

| Layer | Input | Output | Input channel | Output channel |
| --- | --- | --- | --- | --- |
| REBNCONV1 | x | Hx1 | 128 | 64 |
| REBNCONV2 | Hx1 | Hx2 | 64 | 64 |
| REBNCONV3 | Hx2 | Hx3 | 64 | 64 |
| REBNCONV4 | Hx3 | Hx4 | 64 | 64 |
| REBNCONV3_d | Concatenate(Hx4, Hx3) | Hx3_d | 64+64 | 64 |
| REBNCONV2_d | Concatenate(Hx3_d, Hx2) | Hx2_d | 64+64 | 64 |
| REBNCONV1_d | Concatenate(Hx2_d, Hx1) | Hx1_d | 64+64 | 256 |
| REBNCONV_in | x | Hx1_in | 128 | 256 |
| Return | - | Hx1_d+Hx1_in | - | 256 |

REBNCONV (suppose input channel=k, output channe=t)

| Layer | Input | Output | Input channel | Output channel |
| --- | --- | --- | --- | --- |
| Convolution | x | x_conv | k | t |
| Batch normalization | x_conv | x_conv_bn | t | t |
| ReLU | x_conv_bn | x_conv_bn_relu | t | t |
| Return | - | sss | - | t |
