## Supplementary Table S2 for "Deep learning system for brain image-aided diagnosis of multiple major mental disorders"

**Supplementary Table S2** Comparisons of different algorithms on all binary classification tasks, include all possible pairs of diagnoses, and tasks of recognizing specific diagnoses from the others. SVM: support vector machine; XGBoost: extreme gradient boosting, GBDT: gradient boosting decision tree; VGG: 3D visual geometry group network; ResNet: 3D residual network; PHN: patch-based hierarchical network; PHN_linear: PHN with LSTM model replaced by multi-layer perceptron; PHN_noagegen: PHN using only T1 image as input; ACC: accuracy; SEN: sensitivity; SPE: specificity; AUC: area under the ROC curve; F1: F1-score; MCC: Matthews correlation coefficient.

|  | Method | ACC | SEN | SPE | AUC | F1 | MCC |
| --- | --- | --- | --- | --- | --- | --- | --- |
| **BDHC** | Logistic regression | 0.6397±0.0220 | 0.8206±0.0522 | 0.3640±0.0536 | 0.7185±0.0147 | 0.7328±0.0222 | 0.2104±0.0517 |
|  | SVM | 0.6369±0.0152 | 0.7604±0.0483 | 0.4490±0.0641 | 0.7100±0.0161 | 0.7161±0.0180 | 0.2204±0.0351 |
|  | XGBoost | 0.6597±0.0256 | 0.8065±0.0709 | 0.4360±0.0479 | 0.6906±0.0286 | 0.7397±0.0308 | 0.2672±0.0498 |
|  | GBDT | 0.6468±0.0207 | 0.8060±0.0357 | 0.4044±0.0719 | 0.6817±0.0328 | 0.7337±0.0141 | 0.2302±0.0533 |
|  | Random Forest | 0.6910±0.0343 | 0.8826±0.0308 | 0.3990±0.0586 | 0.7220±0.0314 | 0.7753±0.0240 | 0.3277±0.0856 |
|  | VGG | 0.6217±0.0578 | 0.7467±0.0947 | 0.4358±0.1057 | 0.6790±0.0689 | 0.7006±0.0576 | 0.1949±0.1147 |
|  | ResNet | 0.6484±0.0378 | 0.8480±0.0747 | 0.3486±0.0984 | 0.7043±0.0523 | 0.7418±0.0365 | 0.2334±0.0756 |
|  | PHN_linear | 0.8960±0.0131 | 0.8796±0.0236 | 0.9210±0.0164 | 0.9503±0.0142 | 0.9107±0.0124 | 0.7900±0.0239 |
|  | PHN | 0.8921±0.0110 | 0.8871±0.0115 | 0.9083±0.0176 | 0.9557±0.0076 | 0.9080±0.0093 | 0.7801±0.0228 |
| **BDMDD** | Logistic regression | 0.7478±0.0308 | 0.9529±0.0260 | 0.1775±0.1055 | 0.7051±0.0243 | 0.8477±0.0177 | 0.1984±0.1325 |
|  | SVM | 0.7358±0.0021 | 1.0000±0.0000 | 0.0000±0.0000 | 0.6612±0.0345 | 0.8478±0.0014 | 0.0000±0.0000 |
|  | XGBoost | 0.7596±0.0369 | 0.9108±0.0218 | 0.3392±0.0962 | 0.7020±0.0406 | 0.8481±0.0224 | 0.3007±0.1237 |
|  | GBDT | 0.7313±0.0407 | 0.8859±0.0379 | 0.3006±0.0527 | 0.6813±0.0475 | 0.8289±0.0275 | 0.2269±0.1124 |
|  | Random Forest | 0.7615±0.0330 | 0.9375±0.0221 | 0.2713±0.0745 | 0.6921±0.0246 | 0.8526±0.0200 | 0.2844±0.1238 |
|  | VGG | 0.5887±0.0462 | 0.6163±0.0683 | 0.5636±0.0377 | 0.6108±0.0392 | 0.5865±0.0559 | 0.1805±0.0936 |
|  | ResNet | 0.5943±0.0362 | 0.6430±0.0845 | 0.5482±0.0498 | 0.6134±0.0394 | 0.5987±0.0579 | 0.1932±0.0771 |
|  | PHN_linear | 0.8670±0.0153 | 0.8859±0.0136 | 0.8144±0.0337 | 0.9221±0.0104 | 0.9074±0.0108 | 0.6746±0.0371 |
|  | PHN | 0.8963±0.0111 | 0.8985±0.0118 | 0.8900±0.0219 | 0.9295±0.0127 | 0.9272±0.0079 | 0.7521±0.0257 |
| **BDSZ** | Logistic regression | 0.7362±0.0106 | 0.1749±0.0584 | 0.9487±0.0296 | 0.6239±0.0165 | 0.2614±0.0639 | 0.2008±0.0263 |
|  | SVM | 0.7305±0.0121 | 0.0306±0.0612 | 0.9960±0.0079 | 0.5715±0.0411 | 0.0508±0.1017 | 0.0516±0.1032 |
|  | XGBoost | 0.7059±0.0181 | 0.2947±0.0345 | 0.8620±0.0216 | 0.6099±0.0240 | 0.3549±0.0323 | 0.1821±0.0390 |
|  | GBDT | 0.7111±0.0081 | 0.2587±0.0298 | 0.8825±0.0177 | 0.6223±0.0121 | 0.3292±0.0240 | 0.1740±0.0113 |
|  | Random Forest | 0.7270±0.0162 | 0.1805±0.0526 | 0.9346±0.0409 | 0.6042±0.0220 | 0.2616±0.0493 | 0.1830±0.0338 |
|  | VGG | 0.6778±0.0548 | 0.1942±0.0448 | 0.8694±0.0918 | 0.5657±0.0309 | 0.2537±0.0265 | 0.0994±0.0869 |
|  | ResNet | 0.6660±0.0443 | 0.2565±0.0622 | 0.8285±0.0835 | 0.5678±0.0400 | 0.3001±0.0310 | 0.1034±0.0441 |
|  | PHN_linear | 0.6328±0.0216 | 0.5814±0.0279 | 0.6524±0.0260 | 0.6770±0.0287 | 0.4658±0.0194 | 0.2122±0.0366 |
|  | PHN | 0.6752±0.0361 | 0.5929±0.0435 | 0.7058±0.0630 | 0.6833±0.0250 | 0.5020±0.0113 | 0.2783±0.0337 |
| **BDOCD** | Logistic regression | 0.7773±0.0207 | 0.8720±0.0651 | 0.5964±0.0704 | 0.8328±0.0113 | 0.8364±0.0232 | 0.4965±0.0339 |
|  | SVM | 0.7567±0.0271 | 0.8227±0.0582 | 0.6303±0.0733 | 0.8199±0.0143 | 0.8161±0.0229 | 0.4587±0.0659 |
|  | XGBoost | 0.7568±0.0324 | 0.8453±0.0510 | 0.5881±0.0276 | 0.7929±0.0241 | 0.8201±0.0269 | 0.4492±0.0704 |
|  | GBDT | 0.7610±0.0281 | 0.8364±0.0446 | 0.6172±0.0500 | 0.7913±0.0284 | 0.8210±0.0250 | 0.4634±0.0543 |
|  | Random Forest | 0.7937±0.0077 | 0.9117±0.0496 | 0.5701±0.0734 | 0.8445±0.0225 | 0.8529±0.0102 | 0.5301±0.0214 |
|  | VGG | 0.7704±0.0055 | 0.8447±0.0606 | 0.7268±0.0370 | 0.8226±0.0125 | 0.7973±0.0097 | 0.5551±0.0200 |
|  | ResNet | 0.7682±0.0098 | 0.8249±0.0305 | 0.7336±0.0167 | 0.8344±0.0176 | 0.7974±0.0125 | 0.5419±0.0232 |
|  | PHN_linear | 0.8606±0.0215 | 0.8715±0.0238 | 0.8398±0.0359 | 0.9195±0.0071 | 0.8916±0.0170 | 0.6980±0.0473 |
|  | PHN | 0.8784±0.0067 | 0.8817±0.0081 | 0.8717±0.0089 | 0.9204±0.0137 | 0.9051±0.0065 | 0.7374±0.0151 |
| **BDOthers** | Logistic regression | 0.8052±0.0062 | 0.0000±0.0000 | 0.9985±0.0020 | 0.6053±0.0110 | 0.0000±0.0000 | -0.0105±0.0133 |
|  | SVM | 0.8064±0.0064 | 0.0000±0.0000 | 1.0000±0.0000 | 0.4853±0.0226 | 0.0000±0.0000 | 0.0000±0.0000 |
|  | XGBoost | 0.7827±0.0125 | 0.0747±0.0299 | 0.9526±0.0126 | 0.5525±0.0136 | 0.1159±0.0410 | 0.0471±0.0449 |
|  | GBDT | 0.7727±0.0108 | 0.0810±0.0364 | 0.9389±0.0205 | 0.5701±0.0167 | 0.1185±0.0487 | 0.0312±0.0490 |
|  | Random Forest | 0.8016±0.0108 | 0.0228±0.0208 | 0.9885±0.0111 | 0.5650±0.0102 | 0.0407±0.0345 | 0.0358±0.0448 |
|  | VGG | 0.5871±0.0272 | 0.5904±0.0788 | 0.5863±0.0271 | 0.6247±0.0364 | 0.3292±0.0430 | 0.1346±0.0657 |
|  | ResNet | 0.5983±0.0568 | 0.5729±0.0391 | 0.6036±0.0720 | 0.6297±0.0376 | 0.3315±0.0348 | 0.1371±0.0535 |
|  | PHN_linear | 0.6510±0.0256 | 0.6723±0.0263 | 0.6461±0.0344 | 0.7172±0.0124 | 0.4277±0.0233 | 0.2560±0.0288 |
|  | PHN | 0.6803±0.0245 | 0.6524±0.0336 | 0.6871±0.0223 | 0.7253±0.0202 | 0.4418±0.0249 | 0.2765±0.0439 |
| **MDDHC** | Logistic regression | 0.6026±0.0338 | 0.2166±0.1584 | 0.8137±0.1244 | 0.5405±0.0577 | 0.2479±0.1372 | 0.0294±0.0811 |
|  | SVM | 0.5986±0.0433 | 0.2106±0.1258 | 0.8108±0.1240 | 0.4912±0.0638 | 0.2504±0.0998 | 0.0479±0.1087 |
|  | XGBoost | 0.5700±0.0642 | 0.4241±0.1125 | 0.6498±0.1480 | 0.5621±0.0313 | 0.4066±0.0438 | 0.0865±0.1006 |
|  | GBDT | 0.5703±0.0383 | 0.4648±0.1740 | 0.6279±0.1369 | 0.5699±0.0468 | 0.4178±0.0888 | 0.0947±0.0703 |
|  | Random Forest | 0.5821±0.0548 | 0.3259±0.1573 | 0.7223±0.1491 | 0.5869±0.0630 | 0.3343±0.1092 | 0.0632±0.0965 |
|  | VGG | 0.6288±0.0462 | 0.3520±0.0926 | 0.7918±0.0820 | 0.6420±0.0688 | 0.7241±0.0479 | 0.1642±0.0770 |
|  | ResNet | 0.6611±0.0402 | 0.3925±0.0886 | 0.8256±0.0587 | 0.6884±0.0439 | 0.7513±0.0346 | 0.2442±0.0832 |
|  | PHN_linear | 0.6351±0.0588 | 0.6287±0.0826 | 0.6385±0.0506 | 0.6978±0.0589 | 0.5495±0.0717 | 0.2569±0.1222 |
|  | PHN | 0.6866±0.0436 | 0.6701±0.0750 | 0.6956±0.0338 | 0.7191±0.0594 | 0.6014±0.0599 | 0.3529±0.0953 |
| **MDDSZ** | Logistic regression | 0.9002±0.0176 | 0.2471±0.1192 | 0.9890±0.0057 | 0.8297±0.0192 | 0.3622±0.1612 | 0.3771±0.1720 |
|  | SVM | 0.9216±0.0055 | 0.3685±0.0704 | 0.9968±0.0046 | 0.8385±0.0111 | 0.5254±0.0733 | 0.5611±0.0520 |
|  | XGBoost | 0.9057±0.0202 | 0.4506±0.0558 | 0.9678±0.0181 | 0.8492±0.0308 | 0.5368±0.0852 | 0.5001±0.1042 |
|  | GBDT | 0.8925±0.0170 | 0.5366±0.0715 | 0.9410±0.0207 | 0.8251±0.0212 | 0.5454±0.0623 | 0.4881±0.0716 |
|  | Random Forest | 0.9154±0.0046 | 0.4734±0.0622 | 0.9756±0.0110 | 0.8451±0.0128 | 0.5710±0.0366 | 0.5457±0.0326 |
|  | VGG | 0.6300±0.0545 | 0.5266±0.0694 | 0.6751±0.1037 | 0.6256±0.0201 | 0.4649±0.0234 | 0.1988±0.0666 |
|  | ResNet | 0.6172±0.0390 | 0.5736±0.0212 | 0.6365±0.0601 | 0.6496±0.0248 | 0.4782±0.0268 | 0.1970±0.0531 |
|  | PHN_linear | 0.9460±0.0189 | 0.8251±0.0713 | 0.9623±0.0130 | 0.9535±0.0195 | 0.7853±0.0772 | 0.7559±0.0873 |
|  | PHN | 0.9466±0.0246 | 0.8544±0.0528 | 0.9591±0.0260 | 0.9486±0.0149 | 0.7980±0.0823 | 0.7726±0.0914 |
| **MDDOCD** | Logistic regression | 0.7433±0.0346 | 0.5323±0.0987 | 0.8934±0.0776 | 0.7955±0.0238 | 0.6245±0.0656 | 0.4742±0.0827 |
|  | SVM | 0.7501±0.0270 | 0.5141±0.0746 | 0.9148±0.0483 | 0.7859±0.0366 | 0.6243±0.0583 | 0.4834±0.0762 |
|  | XGBoost | 0.7218±0.0466 | 0.6140±0.0962 | 0.8019±0.0868 | 0.8032±0.0282 | 0.6416±0.0642 | 0.4286±0.0961 |
|  | GBDT | 0.7332±0.0284 | 0.6594±0.0422 | 0.7865±0.0375 | 0.7978±0.0392 | 0.6688±0.0378 | 0.4482±0.0593 |
|  | Random Forest | 0.7314±0.0529 | 0.6658±0.0598 | 0.7818±0.1005 | 0.8171±0.0311 | 0.6714±0.0514 | 0.4552±0.1045 |
|  | VGG | 0.7450±0.0060 | 0.5602±0.0864 | 0.8356±0.0405 | 0.7945±0.0145 | 0.8157±0.0028 | 0.4088±0.0268 |
|  | ResNet | 0.7516±0.0232 | 0.5720±0.0827 | 0.8395±0.0266 | 0.8042±0.0198 | 0.8205±0.0153 | 0.4224±0.0554 |
|  | PHN_linear | 0.9373±0.0354 | 0.9300±0.0314 | 0.9425±0.0390 | 0.9740±0.0149 | 0.9226±0.0474 | 0.8702±0.0749 |
|  | PHN | 0.9487±0.0205 | 0.9358±0.0306 | 0.9570±0.0202 | 0.9743±0.0158 | 0.9358±0.0304 | 0.8931±0.0457 |
| **MDDOthers** | Logistic regression | 0.9305±0.0016 | 0.0000±0.0000 | 1.0000±0.0000 | 0.7152±0.0297 | 0.0000±0.0000 | 0.0000±0.0000 |
|  | SVM | 0.9305±0.0016 | 0.0000±0.0000 | 1.0000±0.0000 | 0.5279±0.1821 | 0.0000±0.0000 | 0.0000±0.0000 |
|  | XGBoost | 0.9265±0.0062 | 0.0468±0.0247 | 0.9922±0.0078 | 0.7182±0.0391 | 0.0793±0.0354 | 0.1196±0.0557 |
|  | GBDT | 0.9137±0.0115 | 0.1214±0.0111 | 0.9728±0.0132 | 0.6831±0.0397 | 0.1659±0.0289 | 0.1429±0.0514 |
|  | Random Forest | 0.9309±0.0010 | 0.0175±0.0143 | 0.9991±0.0017 | 0.7191±0.0399 | 0.0333±0.0272 | 0.0820±0.0730 |
|  | VGG | 0.5802±0.0289 | 0.5164±0.0332 | 0.5951±0.0341 | 0.5930±0.0423 | 0.3183±0.0209 | 0.0885±0.0390 |
|  | ResNet | 0.6208±0.0127 | 0.5341±0.0224 | 0.6410±0.0201 | 0.6089±0.0175 | 0.3479±0.0075 | 0.1407±0.0076 |
|  | PHN_linear | 0.8880±0.0147 | 0.7505±0.0816 | 0.8982±0.0136 | 0.9074±0.0206 | 0.4829±0.0559 | 0.4670±0.0636 |
|  | PHN | 0.8799±0.0211 | 0.8319±0.0442 | 0.8835±0.0212 | 0.9129±0.0178 | 0.4941±0.0567 | 0.4918±0.0577 |
| **SZHC** | Logistic regression | 0.8069±0.0156 | 0.9211±0.0161 | 0.3483±0.0394 | 0.7596±0.0372 | 0.8842±0.0103 | 0.3184±0.0429 |
|  | SVM | 0.8006±0.0214 | 0.8935±0.0265 | 0.4277±0.0477 | 0.7502±0.0384 | 0.8775±0.0150 | 0.3434±0.0480 |
|  | XGBoost | 0.8189±0.0209 | 0.9172±0.0132 | 0.4256±0.0648 | 0.7484±0.0529 | 0.8902±0.0129 | 0.3818±0.0671 |
|  | GBDT | 0.8189±0.0168 | 0.9267±0.0139 | 0.3883±0.0960 | 0.7359±0.0540 | 0.8913±0.0097 | 0.3646±0.0690 |
|  | Random Forest | 0.8284±0.0095 | 0.9472±0.0099 | 0.3527±0.0598 | 0.7328±0.0566 | 0.8984±0.0053 | 0.3784±0.0384 |
|  | VGG | 0.7249±0.0966 | 0.8158±0.1257 | 0.3850±0.1472 | 0.6999±0.0959 | 0.8186±0.0806 | 0.2087±0.1646 |
|  | ResNet | 0.7127±0.1039 | 0.7781±0.1298 | 0.4666±0.0726 | 0.7091±0.1138 | 0.8044±0.0912 | 0.2376±0.1388 |
|  | PHN_linear | 0.9502±0.0137 | 0.9487±0.0167 | 0.9558±0.0113 | 0.9830±0.0032 | 0.9681±0.0092 | 0.8582±0.0329 |
|  | PHN | 0.9413±0.0374 | 0.9424±0.0410 | 0.9371±0.0257 | 0.9796±0.0209 | 0.9621±0.0253 | 0.8374±0.0873 |
| **SZOCD** | Logistic regression | 0.8588±0.0210 | 0.9195±0.0300 | 0.5499±0.0611 | 0.8597±0.0215 | 0.9157±0.0134 | 0.4810±0.0708 |
|  | SVM | 0.8430±0.0208 | 0.8990±0.0277 | 0.5575±0.0506 | 0.8338±0.0302 | 0.9053±0.0133 | 0.4468±0.0651 |
|  | XGBoost | 0.8655±0.0218 | 0.9219±0.0199 | 0.5769±0.0621 | 0.8540±0.0199 | 0.9197±0.0132 | 0.5055±0.0793 |
|  | GBDT | 0.8727±0.0158 | 0.9282±0.0140 | 0.5894±0.0523 | 0.8570±0.0135 | 0.9242±0.0091 | 0.5275±0.0686 |
|  | Random Forest | 0.8749±0.0057 | 0.9394±0.0070 | 0.5443±0.0487 | 0.8748±0.0137 | 0.9262±0.0035 | 0.5163±0.0377 |
|  | VGG | 0.8058±0.0332 | 0.8780±0.0528 | 0.4646±0.0661 | 0.7904±0.0378 | 0.8814±0.0227 | 0.3465±0.0863 |
|  | ResNet | 0.8143±0.0308 | 0.8764±0.0544 | 0.5249±0.0645 | 0.8087±0.0262 | 0.8858±0.0218 | 0.3956±0.0572 |
|  | PHN_linear | 0.9578±0.0066 | 0.9684±0.0091 | 0.9050±0.0152 | 0.9743±0.0102 | 0.9746±0.0040 | 0.8515±0.0226 |
|  | PHN | 0.9533±0.0067 | 0.9574±0.0086 | 0.9342±0.0290 | 0.9768±0.0062 | 0.9716±0.0041 | 0.8434±0.0205 |
| **SZOthers** | Logistic regression | 0.6606±0.0183 | 0.7605±0.0445 | 0.5572±0.0543 | 0.7088±0.0198 | 0.6952±0.0188 | 0.3264±0.0363 |
|  | SVM | 0.6566±0.0224 | 0.7818±0.0426 | 0.5269±0.0631 | 0.6995±0.0217 | 0.6987±0.0189 | 0.3212±0.0427 |
|  | XGBoost | 0.6562±0.0173 | 0.7189±0.0552 | 0.5916±0.0480 | 0.7012±0.0134 | 0.6798±0.0236 | 0.3150±0.0366 |
|  | GBDT | 0.6679±0.0213 | 0.7505±0.0622 | 0.5826±0.0524 | 0.7203±0.0180 | 0.6963±0.0270 | 0.3406±0.0457 |
|  | Random Forest | 0.6747±0.0065 | 0.7332±0.0545 | 0.6146±0.0583 | 0.7290±0.0132 | 0.6959±0.0159 | 0.3527±0.0133 |
|  | VGG | 0.6719±0.0166 | 0.7561±0.0352 | 0.6076±0.0418 | 0.7399±0.0143 | 0.6661±0.0142 | 0.3632±0.0290 |
|  | ResNet | 0.6784±0.0142 | 0.7592±0.0290 | 0.6166±0.0099 | 0.7390±0.0152 | 0.6713±0.0179 | 0.3742±0.0321 |
|  | PHN_linear | 0.7735±0.0100 | 0.8494±0.0264 | 0.6945±0.0285 | 0.8289±0.0050 | 0.7925±0.0105 | 0.5522±0.0204 |
|  | PHN | 0.7807±0.0082 | 0.8218±0.0250 | 0.7378±0.0127 | 0.8386±0.0019 | 0.7923±0.0117 | 0.5626±0.0171 |
| **OCDHC** | Logistic regression | 0.6567±0.0387 | 0.7135±0.0325 | 0.6112±0.0526 | 0.7272±0.0392 | 0.6466±0.0433 | 0.3236±0.0697 |
|  | SVM | 0.6483±0.0263 | 0.7503±0.0299 | 0.5660±0.0533 | 0.7125±0.0386 | 0.6526±0.0276 | 0.3182±0.0422 |
|  | XGBoost | 0.6269±0.0269 | 0.6747±0.0582 | 0.5887±0.0639 | 0.6538±0.0415 | 0.6134±0.0361 | 0.2632±0.0453 |
|  | GBDT | 0.6303±0.0302 | 0.6568±0.0616 | 0.6119±0.0627 | 0.6829±0.0581 | 0.6092±0.0391 | 0.2679±0.0602 |
|  | Random Forest | 0.6357±0.0228 | 0.5986±0.0838 | 0.6694±0.0922 | 0.6969±0.0471 | 0.5891±0.0365 | 0.2709±0.0486 |
|  | VGG | 0.6821±0.0282 | 0.7201±0.0407 | 0.6520±0.0513 | 0.7421±0.0244 | 0.6661±0.0264 | 0.3696±0.0534 |
|  | ResNet | 0.6921±0.0196 | 0.7931±0.0461 | 0.6162±0.0352 | 0.7645±0.0131 | 0.6936±0.0243 | 0.4090±0.0403 |
|  | PHN_linear | 0.7466±0.0341 | 0.7960±0.0716 | 0.7060±0.0360 | 0.8152±0.0341 | 0.7323±0.0494 | 0.5012±0.0716 |
|  | PHN | 0.7414±0.0499 | 0.7530±0.0664 | 0.7317±0.0417 | 0.8165±0.0573 | 0.7180±0.0634 | 0.4817±0.1024 |
| **OCDOthers** | Logistic regression | 0.9024±0.0033 | 0.1116±0.0551 | 0.9907±0.0060 | 0.8264±0.0237 | 0.1812±0.0759 | 0.2182±0.0842 |
|  | SVM | 0.8996±0.0080 | 0.0000±0.0000 | 1.0000±0.0000 | 0.5708±0.0633 | 0.0000±0.0000 | 0.0000±0.0000 |
|  | XGBoost | 0.8972±0.0061 | 0.1741±0.0673 | 0.9782±0.0139 | 0.8080±0.0188 | 0.2448±0.0681 | 0.2470±0.0462 |
|  | GBDT | 0.8960±0.0097 | 0.2261±0.0705 | 0.9710±0.0098 | 0.8039±0.0166 | 0.2990±0.0646 | 0.2740±0.0613 |
|  | Random Forest | 0.9020±0.0080 | 0.1020±0.0674 | 0.9915±0.0051 | 0.8297±0.0180 | 0.1644±0.0907 | 0.2026±0.0717 |
|  | VGG | 0.6243±0.0316 | 0.5671±0.0798 | 0.6641±0.0626 | 0.6817±0.0402 | 0.5675±0.0644 | 0.2333±0.0635 |
|  | ResNet | 0.6442±0.0412 | 0.6151±0.1147 | 0.6626±0.0560 | 0.6781±0.0493 | 0.5971±0.0854 | 0.2787±0.0902 |
|  | PHN_linear | 0.8747±0.0091 | 0.8550±0.0196 | 0.8767±0.0108 | 0.9107±0.0147 | 0.5778±0.0197 | 0.5534±0.0191 |
|  | PHN | 0.8863±0.0123 | 0.8786±0.0194 | 0.8871±0.0117 | 0.9285±0.0123 | 0.6078±0.0449 | 0.5871±0.0433 |
| **HCOthers** | Logistic regression | 0.8667±0.0083 | 0.0502±0.0204 | 0.9853±0.0088 | 0.6765±0.0276 | 0.0860±0.0327 | 0.0904±0.0473 |
|  | SVM | 0.8731±0.0032 | 0.0000±0.0000 | 1.0000±0.0000 | 0.6076±0.0155 | 0.0000±0.0000 | 0.0000±0.0000 |
|  | XGBoost | 0.8594±0.0085 | 0.1044±0.0303 | 0.9692±0.0104 | 0.6453±0.0373 | 0.1568±0.0355 | 0.1252±0.0343 |
|  | GBDT | 0.8458±0.0112 | 0.1169±0.0281 | 0.9517±0.0123 | 0.6409±0.0357 | 0.1604±0.0331 | 0.0991±0.0352 |
|  | Random Forest | 0.8667±0.0030 | 0.0410±0.0122 | 0.9867±0.0057 | 0.6105±0.0348 | 0.0718±0.0186 | 0.0738±0.0195 |
|  | VGG | 0.6626±0.0521 | 0.4208±0.0680 | 0.7686±0.0978 | 0.6490±0.0492 | 0.4317±0.0385 | 0.2001±0.0742 |
|  | ResNet | 0.6746±0.0492 | 0.3540±0.0628 | 0.8150±0.0901 | 0.6260±0.0561 | 0.3980±0.0445 | 0.1921±0.0795 |
|  | PHN_linear | 0.8390±0.0111 | 0.7975±0.0408 | 0.8450±0.0138 | 0.8809±0.0220 | 0.5569±0.0260 | 0.5037±0.0296 |
|  | PHN | 0.8361±0.0126 | 0.8325±0.0238 | 0.8367±0.0118 | 0.9001±0.0166 | 0.5636±0.0240 | 0.5160±0.0290 |
