## Supplementary Table S3 for "Deep learning system for brain image-aided diagnosis of multiple major mental disorders"

**Supplementary Table S3** Imaging acquisition parameters details.

|  | **Repetition time (ms)** | **Echo time (ms)** | **Flip angle (°)** | **Field of View (mm^2^)** | **Matrix size** | **Number of slices** | **Slice thickness (mm)** | **Voxel size (mm^3^)** | **Number of samples** |
| --- | --- | --- | --- | --- | --- | --- | --- | --- | --- |
| 1 | 2530 | 3.65 | 7 | 256×256 | 256×256 | 224 | 1 | 1×1×1 | 295 (HC: 116; BD:1; MDD: 93; SZ: 85) |
| 2 | 2530 | 3.65 | 7 | 240×256 | 240×256 | 224 | 1 | 1×1×1 | 1624 (BD: 394; SZ: 1198; OCD: 32) |
| 3 | 2530 | 3.65 | 7 | 176×256 | 176×256 | 224 | 1 | 1×1×1 | 139 (HC: 46; BD: 44; MDD: 48; SZ: 1) |
| 4 | 2300 | 3.5 | 9 | 256×256 | 256×256 | 192 | 1 | 1×1×1 | 259 (HC: 77; SZ: 1; OCD: 181) |
| 5 | 2300 | 2.96 | 9 | 240×256 | 240×256 | 192 | 1 | 1×1×1 | 89 (HC: 34; OCD: 55) |
| 6 | 2300 | 2.96 | 9 | 256×256 | 256×256 | 192 | 1 | 1×1×1 | 84 (HC: 27; BD: 29; MDD: 28) |
