## Supplementary Table S4 for "Deep learning system for brain image-aided diagnosis of multiple major mental disorders"

**Supplementary Table S4** Demographics of four public dataset.

| Dataset | Characteristic | HC | BD | MDD | SZ |
| --- | --- | --- | --- | --- | --- |
| LA5c | Gender (M/F) | 63/57 | 28/20 | - | 11/33 |
|  | Age (Mean±Std) | 31.12±8.62 | 35.56±8.91 | - | 36.25±9.06 |
| COBRE | Gender (M/F) | 50/23 | - | - | 55/14 |
|  | Age (Mean±Std) | 35.95±11.61 | - | - | 37.99±13.67 |
| HBN | Gender (M/F) | 115/93 | - | 20/44 | - |
|  | Age (Mean±Std) | 10.21±3.57 | - | 15.31±2.53 | - |
| SRPBS | Gender (M/F) | 524/402 | 24/14 | 125/112 | 85/60 |
|  | Age (Mean±Std) | 36.16±15.28 | 34.26±9.21 | 42.49±12.25 | 38.04±10.90 |

Abbreviations: LA5c: UCLA Consortium for Neuropsychiatric Phenomics LA5c dataset; COBRE: Centers of Biomedical Research Excellence dataset; HBN: the Healthy Brain Network (HBN) dataset; SRPBS: the Japanese Strategic Research Program for the Promotion of Brain Science (SRPBS) multi-disorder MRI dataset (restricted).

Note: The HBN database contains subjects from release1-9, a total of 2743 valid subjects, including 239 HC and 66 MDD subjects based on consensus diagnostic information. A total of 208 subjects with HC and 64 subjects with MDD had their T1 images quality-controlled by CAT12.6 toolbox (as described in the "Method" section of the main text) and were included in the study.
