## Supplementary Table S5 for "Deep learning system for brain image-aided diagnosis of multiple major mental disorders"

**Supplementary Table S5** Demographics of 323 real-world validation patients.

| Characteristic | BD | MDD | SZ |
| --- | --- | --- | --- |
| Gender (M/F) | 40±43 | 51/76 | 44/69 |
| Age (Mean±Std) | 30.81±15.43 | 40.84±22.09 | 43.44±14.06 |
