## Supplementary Table S6 for "Deep learning system for brain image-aided diagnosis of multiple major mental disorders"

**Supplementary Table S6** Network structure of VGG for comparison experiments.

| Layer | Input | Output | Input channel | Output channel |
| --- | --- | --- | --- | --- |
| Convolution1 | x | x_conv1 | 1 | 8 |
| Batch normalization1 | x_conv1 | x_bn1 | 8 | 8 |
| ReLU1 | x_bn1 | x_relu1 | 8 | 8 |
| Maxpool1 | x_relu1 | x_pool1 | 8 | 8 |
| Convolution2 | x_pool1 | x_conv2 | 8 | 32 |
| Batch normalization2 | x_conv2 | x_bn2 | 32 | 32 |
| ReLU2 | x_bn2 | x_relu2 | 32 | 32 |
| Convolution3 | x_relu2 | x_conv3 | 32 | 32 |
| Batch normalization2 | x_conv3 | x_bn3 | 32 | 32 |
| ReLU3 | x_bn3 | x_relu3 | 32 | 32 |
| Maxpool3 | x_relu3 | x_pool3 | 32 | 32 |
| Convolution4 | x_pool3 | x_conv4 | 32 | 64 |
| Batch normalization4 | x_conv4 | x_bn4 | 64 | 64 |
| ReLU4 | x_bn4 | x_relu4 | 64 | 64 |
| Convolution5 | x_relu4 | x_conv5 | 64 | 64 |
| Batch normalization5 | x_conv5 | x_bn5 | 64 | 64 |
| ReLU5 | x_bn5 | x_relu5 | 64 | 64 |
| Convolution6 | x_relu5 | x_conv6 | 64 | 64 |
| Batch normalization6 | x_conv6 | x_bn6 | 64 | 64 |
| ReLU6 | x_bn6 | x_relu6 | 64 | 64 |
| Maxpool6 | x_relu6 | x_pool6 | 64 | 64 |
| Convolution7 | x_pool6 | x_conv7 | 64 | 128 |
| Batch normalization7 | x_conv7 | x_bn7 | 128 | 128 |
| ReLU7 | x_bn7 | x_relu7 | 128 | 128 |
| Convolution8 | x_relu7 | x_conv8 | 128 | 128 |
| Batch normalization8 | x_conv8 | x_bn8 | 128 | 128 |
| ReLU8 | x_bn8 | x_relu8 | 128 | 128 |
| Convolution9 | x_relu8 | x_conv9 | 128 | 128 |
| Batch normalization9 | x_conv9 | x_bn9 | 128 | 128 |
| ReLU9 | x_bn9 | x_relu9 | 128 | 128 |
| Maxpool9 | x_relu9 | x_pool9 | 128 | 128 |
| Convolution10 | x_pool9 | x_conv10 | 128 | 256 |
| Batch normalization10 | x_conv10 | x_bn10 | 256 | 256 |
| ReLU10 | x_bn10 | x_relu10 | 256 | 256 |
| Convolution11 | x_relu10 | x_conv11 | 256 | 256 |
| Batch normalization11 | x_conv11 | x_bn11 | 256 | 256 |
| ReLU11 | x_bn11 | x_relu11 | 256 | 256 |
| Convolution12 | x_relu11 | x_conv12 | 256 | 256 |
| Batch normalization12 | x_conv12 | x_bn12 | 256 | 256 |
| ReLU12 | x_bn12 | x_relu12 | 256 | 256 |
| AdaptiveMaxpool | x_relu12 | x_pool12 | 256 | 256 |
| Linear1 | x_pool12 | x_linear1 | 256 | 128 |
| Batch normalization13 | x_linear1 | x_bn13 | 128 | 128 |
| ReLU13 | x_bn13 | x_relu13 | 128 | 128 |
| Linear2 | x_relu13 | x_linear2 | 128 | 2 |
| softmax | x_linear2 | out | 2 | 2 |
| Return | - | out | 2 | 2 |

Note: kernel_size is set as 3×3×3 for all Convolution layers; eps is set as 1e-5 and momentum is set as 0.1 for all BatchNorm3D layers.
